# Identification and functional characterisation of a rare *MTTP* variant underlying hereditary non-alcoholic fatty liver disease

**DOI:** 10.1101/2021.07.22.21260356

**Authors:** Jane I. Grove, Peggy Cho Kiu Lo, Nick Shrine, Julian Barwell, Louise V. Wain, Martin D. Tobin, Andrew M. Salter, Neil Bennett, Catherine John, Ioanna Ntalla, Gabriela E. Jones, Christopher P. Neal, Mervyn G. Thomas, Helen Kuht, Pankaj Gupta, Vishwaraj M. Vemala, Allister Grant, Adeolu B. Adewoye, Kotacherry T. Shenoy, Leena K. Balakumaran, Edward J. Hollox, Nicholas R.F. Hannan, Guruprasad P. Aithal

**Author notes:** Corresponding author: Guruprasad P. Aithal, Nottingham Digestive Diseases Centre, Queens Medical Centre Campus, Nottingham NG7 2UH, UK. These authors contributed equally (Jane I. Grove, Peggy Cho Kiu Lo, Nick Shrine). Department of Genetics, Nottingham University Hospitals NHS Trust, Nottingham, UK. Treliske Hospital, Truro, Cornwall, TR1 3LJ, UK. Institute of Inflammation and Ageing, College of Medical and Dental Sciences, University of Birmingham, Birmingham, B15 2TT, UK. Senior Authors (Edward J. Hollox, Nicholas R.F. Hannan, Guruprasad P. Aithal). **Author Contributions** G.P.A. was responsible for conceptualization. G.P.A, J.B, N.R.F.H, A.M.S, L.V.W, and M.D.T. designed the study. J.I.G, P.C.K.L, J.B, N.S, G.P.A, N.R.F.H, E.J.H, N.B, A.G, G.E.J, M.G.T, H.K, A.B.A, P.G and V.M.V completed investigations. G.P.A, N.R.F.H, E.J.H, J.B, J.I.G, M.D.T, and L.V.W supervised the research. J.I.G, P.C.K.L, N.S, N.B, E.J.H, C.J, and I.N. curated data. J.I.G, P.C.K.L, N.S, N.B. formally analysed the data. G.P.A, N.R.F.H, A.M.S, J.I.G, J.B, C.J, I.N, C.P.N, K.T.S, and L.K.B. provided resources. G.P.A, N.R.F.H, E.J.H, K.T.S, M.D.T, and L.V.W. acquired funding. J.I.G, P.C.K.L, N.R.F.H, A.M.S, J.B, L.V.W. and G.P.A. wrote the original draft manuscript and all authors reviewed, edited and approved the final version.

## Abstract

**Background and aims:** Non-alcoholic fatty liver disease (NAFLD) is a complex trait that has a global prevalence estimated as 25%. We aimed to identify the genetic variant underlying a four-generation family with progressive NAFLD leading to cirrhosis, decompensation and development of hepatocellular carcinoma in the absence of common risk factors such as obesity and type 2 diabetes.

**Methods:** Exome sequencing and genome comparisons were used to identify the likely causal variant. We extensively characterised the clinical phenotype and post-prandial metabolic responses of family members with the identified novel variant in comparison to healthy non-carriers and wild type patients with NAFLD. Variant-expressing hepatocyte-like cells (HLCs) were derived from human induced pluripotent stem cells generated from homozygous donor skin fibroblasts. The phenotype was assessed using imaging, targeted RNA analysis and molecular expression arrays.

**Results:** We identified a rare causal variant in *MTTP*, c.1691T>C p.I564T (rs745447480) encoding microsomal triglyceride transfer protein (MTP) associated with progressive non-alcoholic fatty liver disease, unrelated to metabolic syndrome. Although other described mutations in MTTP cause abetalipoproteinemia, neither homozygotes nor heterozygotes exhibited characteristic manifestations of this severe disease. HLCs derived from a homozygote donor had lower lipoprotein ApoB secretion, compared to wild type cells. Cytoplasmic triglyceride accumulation in HLCs triggered endoplasmic reticulum stress, secretion of pro-inflammatory mediators and production of reactive oxygen species.

**Conclusion:** We have identified and characterized a rare causal variant in *MTTP* and homozygosity for *MTTP* p.I564T is associated with progressive NAFLD without any other manifestations of abetalipoproteinemia.

## Introduction

Non-alcoholic fatty liver disease (NAFLD) is a complex trait encompassing a spectrum of accumulation of triglyceride-rich lipid droplets within the hepatocytes (steatosis), non-alcoholic steatohepatitis (NASH; having ballooning degeneration and inflammatory cell infiltration), varying degree and pattern of fibrosis leading to cirrhosis and its decompensation, as well as hepatocellular carcinoma (HCC). With rising incidence of obesity and type 2 diabetes, NAFLD is now the most common chronic liver disease with an estimated 25% population prevalence globally.^1^

Genome-wide association studies (GWAS) have identified a number of genetic risk variants for NAFLD including *PNPLA3* p.I148M (c.444C>G, rs738409) and *TM6SF2* p.E167K (c.449C>T, rs58542926), both of which have robust associations with disease phenotypes via functional pathobiological pathways.^2, 3^ Accretion of Patatin-like phospholipase domain-containing protein 3, PNPLA3, 148M on lipid droplets sequesters coactivators, resulting in reduced lipolysis and lipophagy and transmembrane protein 6 superfamily member 2, TM6SF2, 167K variant impairs VLDL lipidation; accumulation of triglycerides in both contexts is associated with progressive liver disease.^2^

The putative role of microsomal transfer protein (MTP), a lipid transfer protein localized in the endoplasmic reticulum of hepatocytes involved in lipidation and assembly of ApoB containing lipoproteins in the development of NAFLD, has been investigated and *MTTP* variants have been linked with susceptibility to NAFLD.^4–6^ Rare, loss-of-function mutations in *MTTP* can result in the recessive disorder abetalipoproteinemia (ABL)^7^ (OMIM:200100), where MTP deficiency causes defective lipoprotein biosynthesis having multiple severe effects including liver steatosis and fibrosis.^8–10^ However, hereditary progressive NAFLD associated with a *MTTP* variant, without any manifestations of abetalipoproteinemia, has not been previously described.

Here we have clinically characterised a large four generation family found to have a rare *MTTP* variant resulting in progressive NAFLD, with consequent cirrhosis, liver failure and hepatocellular carcinoma in homozygotes. We evaluated post-prandial metabolic responses in carriers of the novel *MTTP* p.I564T variant compared to non-carriers. We used hepatocyte-like cells (HLCs) derived from human induced pluripotent stem cells (hiPSCs) generated from donor skin fibroblasts from carriers and non-carriers of the *MTTP* variant, as a stable reproducible model for understanding the effect of the variant on the cellular phenotype. This has enabled us to understand how this can drive steatosis and NAFLD and therefore linking genotype to phenotype in hereditary NAFLD.

We show that the I564T variant is associated with increased sequestration of lipids in cultured *MTTP^(VAR/VAR)^* HLCs derived from hiPSCs. These HLCs have elevated endoplasmic reticulum (ER) stress, activated pro-inflammatory signalling pathways including NFκB, and secrete pro-inflammatory mediators. This coincides with reduced levels of MTP and apolipoprotein B-100 (ApoB-100), increased production of reactive oxygen species (ROS) and superoxide, and alterations to mitochondrial respiration. This confirms the role of functional genetic variants in maintaining hepatic lipid homeostasis via lipoprotein secretion. This model also provides important insights into how chronic hepatic steatosis can progress to hepatocyte dysfunction, inflammation, cancer and eventually organ failure.

## Materials and Methods

### Patient Investigations

Clinical investigations followed standard clinical care and included 6-month follow-up as required. Family members were screened at joint hepatology-genetics clinics within NHS Trusts between 2013-2020.

### Human Samples

The clinical studies were reviewed by the National Research Ethics Service (Genetics of Rare Inherited Disorders (GRID): East Midlands Northampton Committee (Ref 12/EM/0262); meal-response study: North-East Committee (Ref 16/NE/0251) and approved by the Health Research Authority. Studies were conducted according to the Declaration of Helsinki (Hong Kong Amendment) and Good Clinical Practice (European guidelines). All participants provided written informed consent. Family members have reviewed the manuscript and consent to publication.

### Meal-response study

Participants were recruited to the study at Queens Medical Centre, Nottingham University Hospitals between 1/11/2016 and 1/6/2017. Fasting and post-prandial blood samples were immediately processed to obtain plasma and serum for biomarker and metabolite analyses. Lipoprotein fractions were prepared within 12h and other samples were stored at -80°C and analysed at the University of Nottingham Metabolic Analysis Facility to quantify insulin, glucose, cholesterol, triglycerides, and free fatty acids. Skin biopsy material was collected into culture media and immediately processed for growth.

Standard meals were provided. Breakfast was 70g Kelloggs Cornflakes, 300ml Tesco British Whole Milk with 35g Nestle Lido full cream milk powder (total: 633kcals; 21.3g fat; 84.9g carbohydrate; 23.5g protein; 2.1g fibre; 1.5g salt). Lunch was Sainsbury’s Indian Vegetable Biryani (500g) meal and milkshake containing 200ml Tesco whole milk, 20ml Tesco fresh double cream and Kelly’s clotted cream vanilla ice cream (total: 1134kcals; 58.7g fat; 119.9g carbohydrate; 23.9g protein; 15.7g fibre; 1.77g salt).

### Genetic Variant Identification

DNA was derived from blood except the 3 EXCEED study samples^11^ which were derived from saliva. Whole exome sequencing (single batch with 3 replicates) mean depth of coverage was 42-66×. Exome enrichment was done in 3 batches using NimbleGen SeqCap EZ Exome v3.0 (64Mb). Samples were sequenced using 100bp paired-end sequencing on the Illumina HiSeq2000 (each sample sequenced in 2 lanes) and validated by Sanger sequencing. Both NAFLD cases and controls were present in each pair of lanes and in each batch to minimise confounding.

After alignment using BWA 0.7.5^12^, reads were cleaned with Picard v1.93 (synchronise mate-pair information; http://broadinstitute.github.io/picard/) and samtools v1.1^13^ (PCR duplicates removed, sorted and indexed); local realignment around indels and recalibration of quality scores was done with GATK v3.2-2^14^. Variant calling was done using both GATK v3.2-2 HaplotypeCaller and samtools v1.1 mpileup to obtain a consensus. Concordance between the two methods was 96.8%. Concordance between the 3 pairs of replicate samples was 98.4-98.6%.

First, we identified variants that were not called in any of the 9 control samples. This identified 8,835 SNPs unique to one or more NAFLD cases. We next excluded any SNPs that were also present in the following datasets: 1000 genomes phase I high confidence SNPs (https://www.internationalgenome.org), 1000 Genomes Project May 2013 release, 1000 genomes Illumina OMNI 2.5 SNP array (ftp://ftp.1000genomes.ebi.ac.uk/vol1/ftp/release/20130502/supporting/hd_genotype_chip/), dbSNP version 138 and 142 (https://www.ncbi.nlm.nih.gov/snp/) HapMap3 (https://www.sanger.ac.uk/resources/downloads/human/hapmap3.html), NHLBI exomes (March 2015; https://evs.gs.washington.edu/EVS/) and whole exome sequencing of 125 South Asian samples^15^. We restricted the remaining 1024 SNPs to those annotated as ‘exonic’ using ANNOVAR^16^ (November 2014 version) which left 434. Of those, Variant Effect Predictor^17^ annotated 159 SNPs as ‘deleterious’ by SIFT^18^, ‘probably damaging’ or ‘potentially damaging’ by PolyPhen-2^19^, or had a CADD^20^ Phred scaled score≥20 or GWAVA^21^ score >0.5. Segregation with disease was assessed by overlaying the variant genotypes on the pedigree.

A missense variant in *MTTP* seen in all 12 affected individuals (6 heterozygotes and 6 homozygotes) was the only variant that fully segregated with disease. Two additional variants were found as heterozygotes: N1484S in *NOTCH1* NC_000009.11:g.139399897T>C (10 cases) and G796R in *EPB41L1* NC_000020.10:g.34807716G>C (11 cases).

Variant modelling based on Protein Data Bank sequence 617S^22^ was completed using PyMOL v2.5.0 (Schrodinger Inc).

### Genotyping

Genotype validation was done using Sanger sequencing for *MTTP* alleles (Source Bioscience Ltd) or PCR restriction fragment analysis using primers listed in Supplemental Methods.

### Metabolite analyses

Plasma lipoproteins were separated by sequential non-equilibrium density-gradient ultracentrifugation^23^. EDTA plasma was pipetted into quick-seal ultracentrifuge tubes (Beckman Coulter) and topped up with 1.006g/ml potassium bromide (KBr). Sealed tubes were centrifuged in a Beckman Optima ultracentrifuge XL-70 under vacuum at 12°C for 20min at 12,000rpm. Afterwards, each tube was opened using a tube slicer (Kontron). The top chylomicron layer was taken and made up to 2ml with KBr solution and stored at -20°C. The lower lipoprotein layer was then transferred to fresh ultracentrifuge tubes and centrifuged for 16h at 39,000rpm with full acceleration and no break. Following ultracentrifugation the top VLDL layer was removed, this and the lower layer were separately stored at -20°C.

Blood metabolites were quantified using calibrated Horiba auto-analyser and reagents following standard manufacturer validated protocols (Horiba ABX). Serum ApoB was determined by turbidimetry using ABX Pentra Apo reagent. Serum cholesterol was quantified using cholesterol BP diagnostic reagent, and colorimetry with HDL Direct CP and LDL Direct CP reagents used for LDL and HDL-cholesterol. Glucose was quantified in plasma by colorimetry using Glucose PAP CP reagent. Colorimetry with Triglycerides CP reagent was used to quantify serum triglycerides. Plasma free fatty acids were measured using Wako NEFA C enzymatic colour test method (Wako Chemicals GmbH). Serum insulin was quantified using Human Insulin specific RAI kit (Millipore).

### Isolation and culture of human dermal fibroblasts

Two 2mm skin punch biopsies were obtained from study participant 1, a healthy male, and participant J, (homozygous for rs745447480 I564T). Primary human dermal fibroblasts were established via by explants culture in DMEM medium supplemented with 2% Antibiotic-Antimycotic; 10% FBS; 1% GlutaMAX; 1% NEAA and 1% penicillin/streptomycin. Fibroblast medium was refreshed every 2-3 days, cells were split at 1:3-1:6 using 0.25% Trypsin-EDTA for 3min at 37°C when 80% confluent. All reagents were from Gibco (Thermo Fisher). All cell lines tested negative for mycoplasma contamination using the EZ-PCR Mycoplasma Test Kit (Biological Industries) prior to reprogramming.

### Fibroblast reprogramming, hiPSC maintenance and differentiation

Human skin fibroblasts were reprogrammed using CytoTune iPS 2.0 Sendai Reprogramming Kit (Invitrogen, Thermo Fisher) in accordance with the manufacturer’s guidelines. Mesoderm and ectoderm differentiation of hiPSCs was as described previously^24, 25^. All cells were differentiated into hepatocytes as described previously^26^.

### Karyotyping

30 metaphase spreads from exponentially growing hiPSC cultures were analysed by conventional karyotyping^27^ (Nottingham University Hospitals).

### Protein analyses

Human Apolipoprotein B-100 (apoB-100) was determined in culture supernatants by ELISA (Sigma-Aldrich) according to the manufacturer’s recommendations (in duplicate). Human NFκB Pathway, Phospho-Kinase and XL Cytokine Array Kits were used as specified by the manufacturer (R&D Systems).

### Imaging and detection of cellular and mitochondrial ROS

Cells were stained with Nile Red, Hoechst, DAPI or antibodies listed in Supplemental Methods. Mitochondrial content of HLCs were visualised using 100nM MitoTracker green FM or MitoTracker deep red FM and intracellular reactive oxygen species (ROS) and mitochondrial superoxide production were assessed using 2.5μM CellROX green or 2.5μM MitoSox Red following manufacturer’s guidance (Invitrogen).

### Mitochondrial respiration analysis

To assess mitochondrial respiration, culture medium was replaced with 200µl Seahorse XF base medium supplemented with 10mM glucose, 1mM sodium pyruvate and 2mM L-glutamine at 37°C without CO_2_ for 1h prior to measurements using the Seahorse XF96 analyser (Seahorse Bioscience, USA). Mitochondria stress tests were performed as recommended by the manufacturer (Agilent), oxygen consumption rate (OCR) was measured while injecting oligomycin (1.5μM), FCCP (0.4μM), rotenone (1μM) and XF base medium. OCR values were normalised by the number of viable cells counted with DAPI.

### RT-qPCR Gene expression analysis

Quantitative real-time PCR (qPCR) was carried out as described previously^28^. Fold changes in expression were calculated using comparative ΔΔCt method standardised against the housekeeping gene PBGD, data were shown as mean of Ct values ± standard error of mean (SEM).

### RNA sequencing

RNA sequencing and bioinformatics analysis were performed at the Babraham Institute (Cambridge, UK). Sequencing was done using the Illumina HiSeq2500 system (depth = 30 million). Reads were mapped to Ensembl GRCh38.p10 using Hisat2 (v2.1.0). Analysis used SeqMonk (1.46.0) software, with read counts determined using RNA-Seq pipeline and differential expression analysis using DESeq2 (1.28.1) package. Data was trimmed with Trim Galore v0.6.2 (default parameters), aligned to the GRCh38 using Hisat2 v2.1.0 (option “--sp 1000,1000” to prevent soft-clipping) then seeded with introns from gene models from Ensembl v87. Alignments with a MAPQ score of <20 were discarded.

Per gene expression was quantitated against gene models from Ensembl v97 counting read overlaps to any exon of each gene. Only alignments on the opposite strand to the gene being measured were counted. For normalised expression visualisation log2 Reads per million reads of library (log2RPM) values were calculated, and corrected using size factor normalisation based on genes with replicate measurements.

An initial set of differentially expressed genes was calculated from raw counts using the DESeq2 package. Genes with a FDR of <0.05 were retained. This list was further filtered using an expression normalised fold change z-score, again with a cut-off of FDR <0.05.

### Statistical analysis

Data are shown as means ±standard error of the mean unless stated otherwise. Statistical analyses were performed using Graph Pad Prism version 8 (La Jolla, CA, USA) software. One-way ANOVA followed by Dunnett’s multiple comparison test were used to compare data from samples grouped by a single factor. Two-way ANOVA followed by Sidak’s post hoc test was used to compare data grouped by two factors. All authors had access to the study data and reviewed and approved the final manuscript.

## Results

### Clinical presentation of family

A four-generation family with recent South Asian ancestry was referred to the Clinical Genetics Department for genetic counselling after three individuals from the same generation developed hepatocellular carcinoma (HCC). One parent of that generation (B in Table 1) presented with NAFLD symptoms and was diagnosed with cirrhosis; the other parent (A) had no history of NAFLD or metabolic syndrome. Clinical investigations found all their children had NAFLD diagnosis as adults with progression to NASH, cirrhosis (in seven) and HCC (in four) suggestive of a high conversion rate between NAFLD to cirrhosis and hepatocellular carcinoma. Only one of the affected individuals had a body mass index >30 and instances of type 2 diabetes, hypertension or hyperlipidaemia within the family were not linked with the presence or severity of disease (Table 1).

**Table 1.** Clinical characteristics of family members.

| Family Member<sup>1</sup> | MTTP genotype: MTP residue 564 (T is variant) | Sex | Liver Disease Diagnosis | T2D | BMI | Hypertension | Liver biochemistry & lipid blood analyses at diagnosis | Other subsequent investigations, treatments and comorbidities |
| --- | --- | --- | --- | --- | --- | --- | --- | --- |
| B | I T | F | Cirrhosis | x | <30 | ✓ | Normal liver biochemistry, Normal lipids |  |
| C | T T | F | HCC, cirrhosis | ✓ | <30 | ✓ | Elevated ALT, Normal lipids, |  |
| D | T T | M | Cirrhosis | x | <30 | ✓ | Normal lipids |  |
| E | I T | M | FL | ✓ | ≥30 | ✓ | Normal liver biochemistry, Normal lipids | Stable for >10y: TE<7kPa; CAP>302; Normal liver biochemistry |
| F | T T | M | Cirrhosis | x | <30 | ✓ | Normal liver biochemistry, Normal lipids |  |
| G | (T*)<sup>2</sup> | M | HCC, cirrhosis | x | <30 | x | Normal lipids |  |
| H | T T | M | HCC, cirrhosis, | x | <30 | ✓ | Normal lipids | Liver transplant. |
| I | I I | F | FL | x | <30 | x |  | Recovered (TE<7kPa; CAP<302), Normal liver biochemistry |
| J | T T | F | Cirrhosis, HCC | x | <30 | ✓ | Normal liver biochemistry, Normal lipids | Liver transplant. Recovered (TE<7kPa; CAP<302), Normal liver biochemistry |
| K | I T | F | NASH (USS) (TE-CAP>302) | x | <30 | ✓ | Elevated ALT; Normal lipids; | Recovered (TE<7kPa; CAP<302), Normal liver biochemistry |
| L | T T | F | Cirrhosis (USS) | x | <30 | ✓ | Elevated ALT<br>Normal lipids | Stable (TE>7kPa; CAP<302), |
| M | I I | M | Healthy | x | <30 | x | Normal lipids | TE<7kPa; CAP<302, Normal liver biochemistry; |
| N | I T | M | NASH (USS) | x | <30 | x | TE<7kPa CAP>302; lipids and ALT elevated | Stable |
| O | I T | M | FL (USS) | x | <30 | x | lipids elevated | Recovered (TE<7kPa; CAP>302), Normal liver biochemistry |
| Q | I T | M | FL (USS) | x | <30 | x | Normal lipids | Recovered (TE<7kPa; CAP<302), Normal liver biochemistry |

<sup>1</sup> Letters refer to individuals from family indicated in illustrative pedigree available from authors; *MTTP* p.I564T novel variant (variant allele nucleotide C results in threonine residue 564 (T)); <sup>2</sup> likely homozygote CC. F: female; M: male; USS: abdominal ultrasonographic steatosis score; HCC: hepatocellular carcinoma; NASH: non-alcoholic steatohepatitis; FL: fatty liver; Bx: biopsy; T2D: Type 2 diabetes mellitus; ALT: alanine transaminase U/L; TE: transient elastography; CAP: Controlled Attenuation Parameter dB/m; ✓ present; x absent. Further clinical details are available from the authors.

### Identification of rare MTTP variant allele associated with diagnosis

Functional variants which were unique to affected family members were identified by whole exome sequencing of twelve affected individuals (B, C, D, E, F, H, J, K, L, N, O, P in Table 1) by comparison with nine unaffected South Asian controls (including spouses of four affected individuals), three unrelated population-based participants from the EXCEED study^11^, and two unrelated South Asian individuals with cholangiocarcinoma) and databases of genetic variation identified in the general population. We identified a missense variant: genomic NC_000004.12:g.99608899T>C, NM_000253.2:c.1691T>C, protein NP_000244.2:p.Ile564Thr, abbreviated to I564T, in *MTTP* that was unique to affected family members (Figure 1) and fully segregated with disease phenotype in those individuals analysed. All six homozygous individuals developed cirrhosis and three also developed hepatocellular carcinoma, while some heterozygotes had no diagnosed disease (Table 1). Presence of both heterozygotes and homozygotes for the rare allele in among the siblings in the family implies that individual A must also have carried at least one copy of the variant allele. The presence of fatty liver in wild type individual I is suggested to be incidental, relating to lifestyle factors.

**Figure 1.**
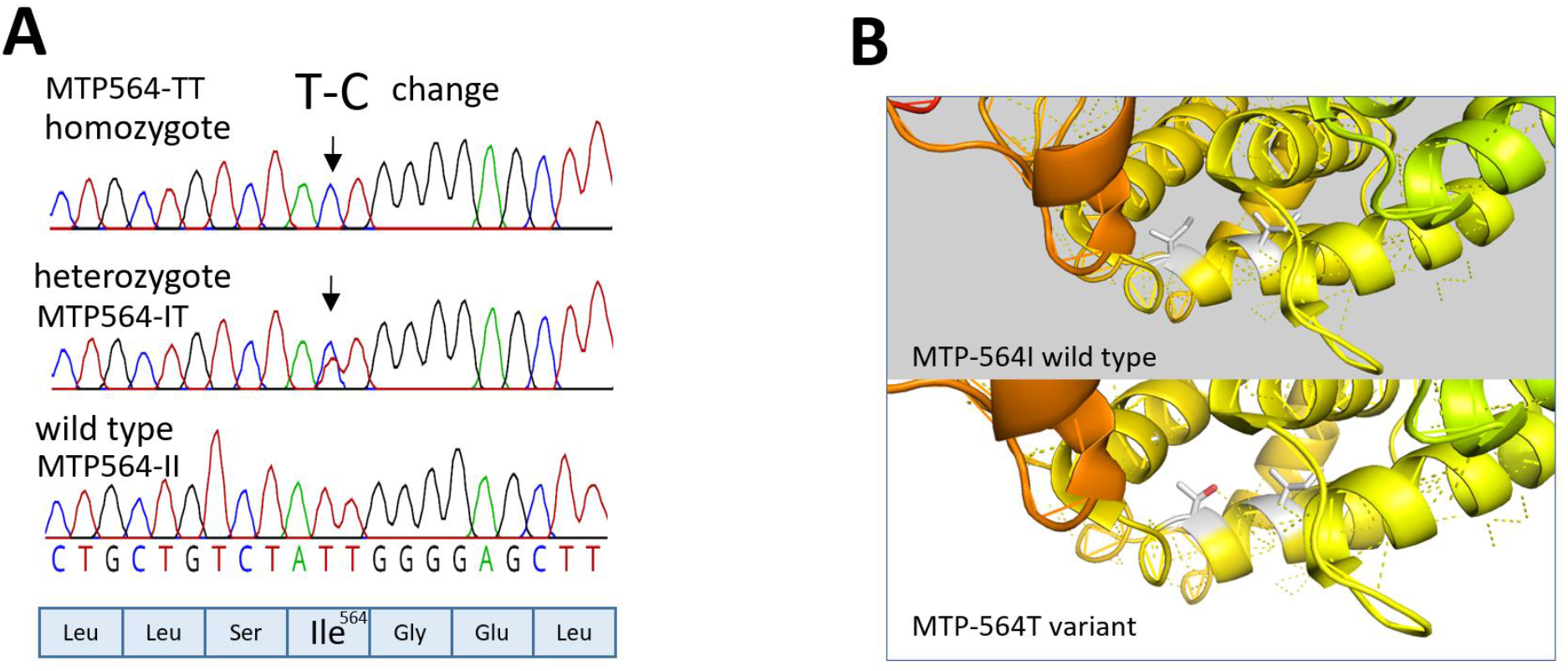
Identification of a pathogenic variant in large family with non-alcoholic fatty liver disease. **(A)** DNA change resulting in I564T amino acid variant identified in homozygous and heterozygous family members. This is annotated as rs745447480 in the NCBI database. **(B)** MTP Model derived in PyMol showing position of substitution I564T based on PDB structure 617S from PDB (Biterova et al., 2019). The side chain at position 564 and 560 is shown as a stick. The favoured rotamer (91.3% probability) for Threonine 564 is shown which is proposed to form a hydrogen bond to Isoleucine 560 causing a minor change in the helix structure.

This I564T variant has been previously described in combination with a second rare variant (IVS1+1G>C), manifesting as severe fatty liver in an atypical case of ABL in Japan^29^ but was reported to have ‘mild effect’ in the mother carrying I564T alone. The I564T variant is described in the NCBI database^30^ as rs745447480 with allele frequency <1.6 x 10^5^ (gnomAD exomes v2.1.1) present in 4 European cases. Other family members were subsequently tested for this variant and clinical features of ABL^7^ were investigated (Table 1 & Supplementary Table 1). Their genotype for other common functional variants at loci in *MTTP* (rs745447480, rs3816873, rs2306985), *PNPLA3* (rs738409) and *TM6SF2* (rs58542926) associated with NAFLD was also determined (Supplementary Table 2). None of the 83 NAFLD patients tested from the Trivandrum Indian cohort^31^ had the *MTTP* p.I564T variant allele.

### Postprandial responses in affected individuals

The *MTTP* p.I564T variant is predicted to have a potential impact on MTP function (PolyPhen=1; GERP=5.120; CADD=20.4, Mutation Assessor=0.76, REVEL=0.519^32^) which we assessed through investigation of metabolic responses to fat consumption. Family members were invited and responses in five participants were compared to that in age and gender matched healthy volunteers and NAFLD patients (Figure 2A; Supplementary Table 3). Levels of ApoB (Figure 2B), the protein constituent assembled into chylomicron and very low density lipoproteins (VLDL) via the activity of MTP, was substantially lower in the MTP564-TT homozygote F. Levels in other participants including two MTP564-IT heterozygotes (participants K and Q) and the MTP564-TT liver-transplant recipient J, was within the normal clinical range (Figure 2B). Subsequent testing of 6 further heterozygotes also showed levels within the normal range (Supplementary Table 1) while testing of a stored pre-transplant serum sample from individual J and clinical data revealed that levels were also substantially lower before receiving a replacement liver where the gene is likely restored (Table 1).

**Figure 2.**
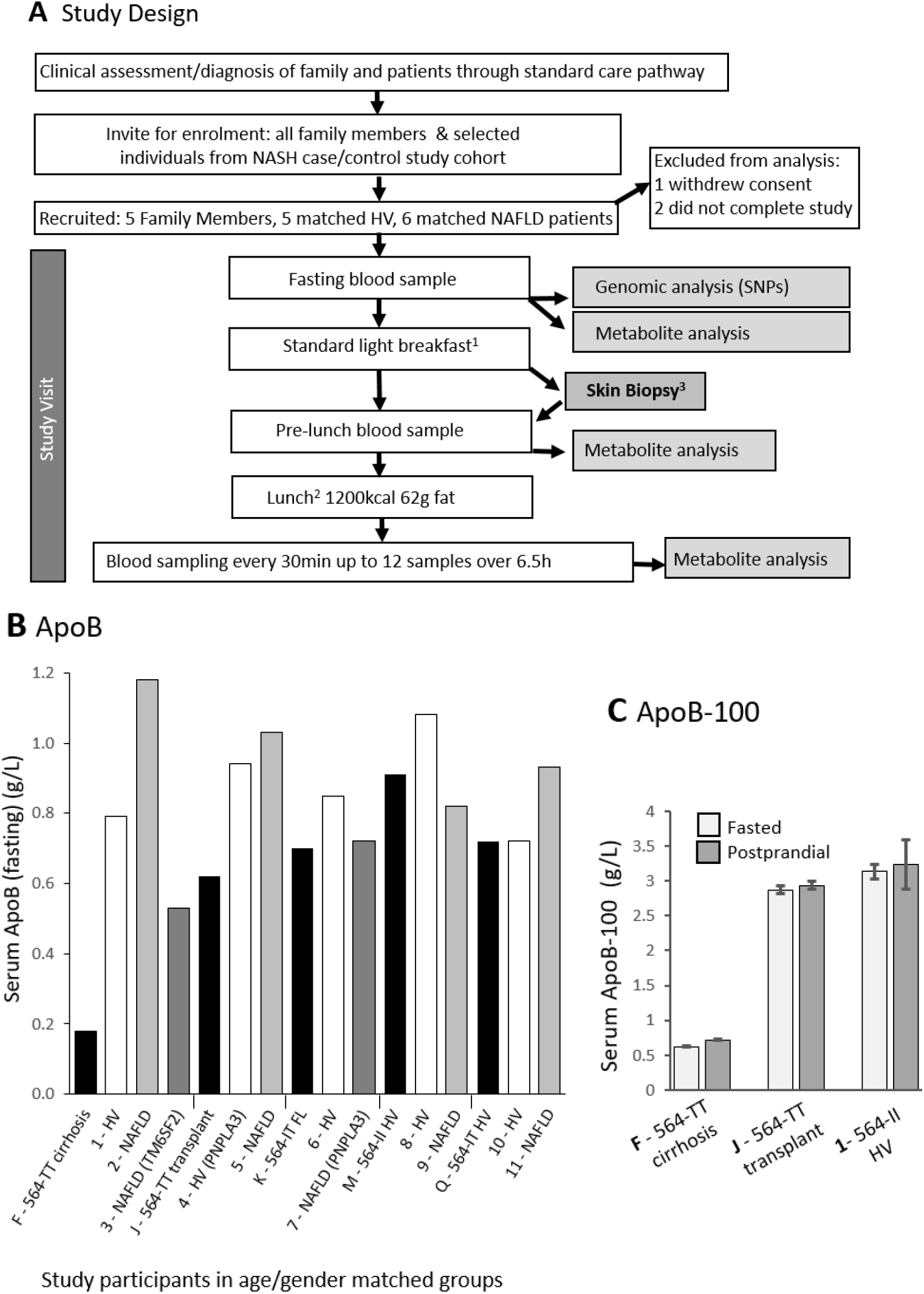
Meal-response study to investigate metabolism in family members and matched controls. **(A)** Study Design. ^1^ milk and cornflakes. ^2^ 80-175 min between breakfast started and pre-lunch sample.^3^ from participants J and 1 used in tissue culture to derive cell lines. (Participants J, K and 1 did not fully consume the test meal). **(B)** ApoB levels in study participants (clinical normal range=0.4-1.25g/L). Family members who participated (black bars) were: F and J being MTP564-TT homozygous, K and Q being MTP564-IT heterozygous and M being MTP564-II (Supplementary Tables 1 and 2). Participants are grouped according to age and gender matched to each family member (white bars are healthy volunteers (HV); grey bars are NAFLD patients). Genes are shown in parentheses where participant is homozygous for the effect allele (PNPLA3 rs738409; TM6SF2 rs58542926). NAFLD= non-alcoholic fatty liver disease; FL=Fatty Liver. **(C)** ApoB-100 levels in serum from study participants fasted and approximately 3 hours after eating.

We also compared levels of serum ApoB-100, the isoform associated with VLDL, in the MTP564-TT homozygotes F and J, with that of wild type MTP564-II control, participant 1 (Supplementary Table 3), to support the proposal that expression of the variant form in the liver results in reduced VLDL secretion (Figure 2C). Levels of total cholesterol both before and after the meal were noticeably lower in the MTP564-TT homozygote F (Supplementary Figure 1) compared to all other study participants due to only very low levels of HDL-cholesterol being present (<0.4mmol/L). Clinical data confirms this observation and the same phenotype was noted in another MTP564-TT homozygote (H), although levels in homozygote J pre-transplant, were normal. Participant F also reported post-prandial gastrointestinal discomfort and diarrhoea.

The levels of circulating triglyceride for the *MTTP* homozygote F were lower than in the healthy and disease controls (participants 1 and 2 in Figure 3A) but were similar to levels in participant 3 who possessed two variant alleles for *TM6SF2* rs58542926 (TM6SF2-KK). This TM6SF2 protein is also involved in VLDL lipoprotein secretion and the uncommon variant associated with impaired function and decreased serum LDL-cholesterol.^33, 34^ In contrast, the MTP564-TT homozygote with a liver transplant showed a similar triglyceride response to her two matched controls (Figure 3B). Investigation of VLDL and chylomicron lipoprotein associated triglycerides revealed that both components were again lowered in participant F (MTP564-TT) and 3 (TM6SF2-KK) compared to the controls (Figure 3, C and E) while the transplanted MTP564-TT, J, had similar levels to her matched controls (Figure 3, D and F). The other family members had no notable defects in lipoprotein triglycerides (Supplementary Figure 2). Furthermore, VLDL-cholesterol levels were similarly blunted in participants F (MTP564-TT) and 3 (TM6SF2-KK) but not in other family members (Supplementary Figure 3) suggesting that transplant hepatocytes and MTP564-IT heterozygote hepatocytes are functioning effectively in VLDL secretion.

**Figure 3.**
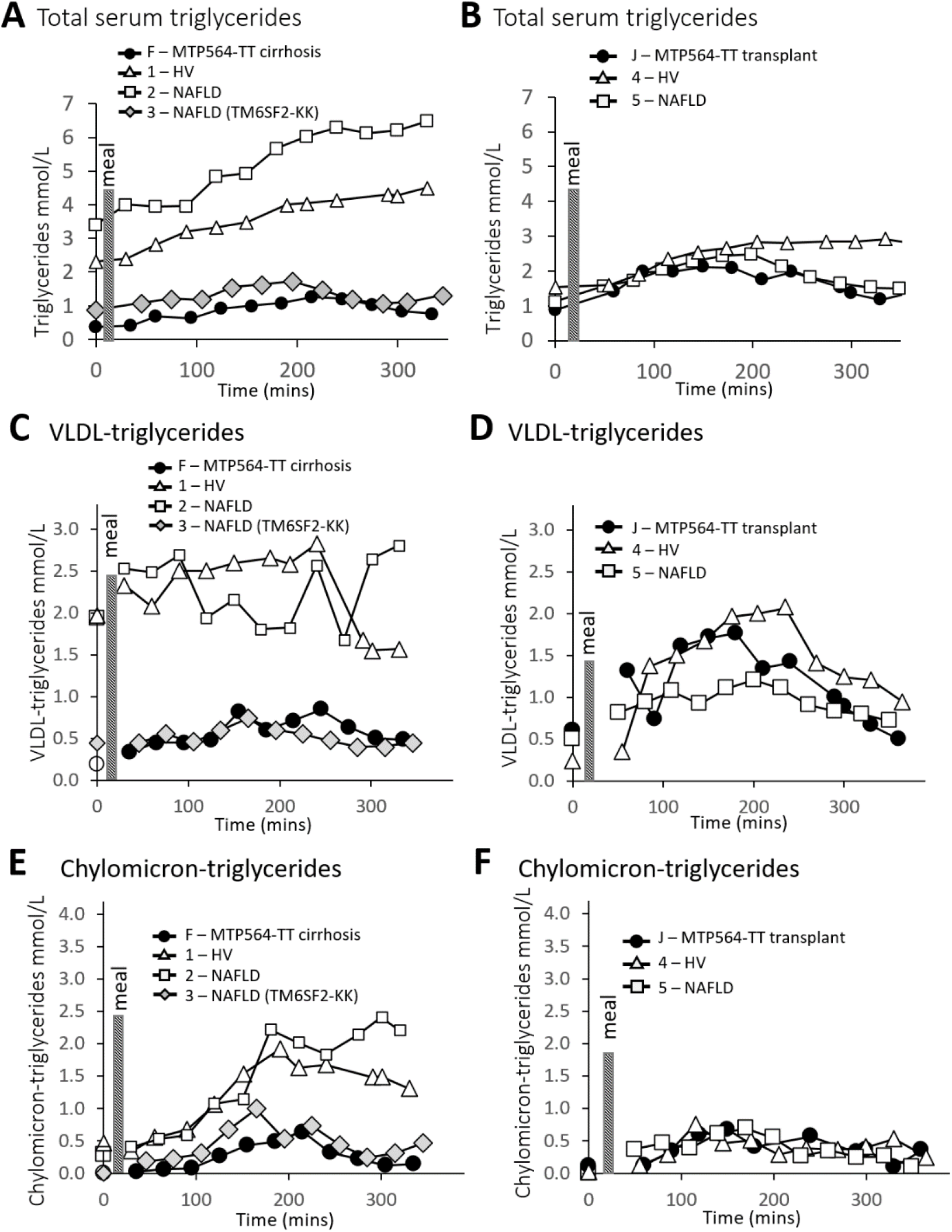
Triglyceride levels in study participants. **(A)** Total serum triglycerides in MTP564-TT family member F with cirrhosis and matched control participants: 1, healthy volunteer (HV) and NAFLD patients 2 and 3. **(B)** Total serum triglycerides in MTP564-TT family member J with liver transplant and matched control participants: 3 (HV) and 4 (NAFLD patient). **(C)** VLDL-triglyceride in MTP564-TT family member F with cirrhosis and matched control participants: 1 (HV), 2 and 3 (NAFLD patients). **(D)** VLDL-triglyceride in MTP564-TT family member J with liver transplant and matched control participants: 3 (HV) and 4 (NAFLD patient). **(E)** Chylomicron-triglyceride in MTP564-TT family member F with cirrhosis and matched control participants: 1 (HV), 2 and 3 (NAFLD patients). **(F)** Chylomicron-triglyceride in MTP564-TT family member J with liver transplant and matched control participants: 4 (HV) and 5 (NAFLD patient). TM6SF2 (rs58542926) variant homozygote (128-KK) is indicated. HV=healthy volunteer; NAFLD=non-alcoholic fatty liver disease.

Of note, lipoprotein-associated lipid levels are also lower in participant 7, a NAFLD patient who is homozygous for the *PNPLA3* rs738409 variant (PNPLA3-MM) which has been linked to a relative reduction in large VLDL secretion^35^ (Supplementary Figures 2A and 3F). Circulating free fatty acids, glucose and insulin levels in the family members were unremarkable (Supplementary Figures 1C, 1D and 4).

### Generation of wild-type and MTTP^(VAR/VAR)^ hiPSCs for disease modelling

To elucidate the mechanisms driving hepatic steatosis in homozygous MTP564-TT patients, we generated hiPSCs and differentiated them into hepatocytes termed *MTTP^(^*^VAR/VAR)^, to create an in vitro model of the *MTTP* variant (p.I564T) (Supplementary Figures 5, 6, and 7). Skin biopsies from participant 1, MTP564-II, and family member J, MTP564-TT, were dissected to remove subcutaneous fat and cultured in fibroblast media for approximately 10 days until fibroblasts began emerging from the biopsy forming a monolayer. Fibroblasts were expanded up to passage 4 and then reprogrammed into hiPSCs: namely *MTTP^(WT/WT)^*, and *MTTP^(^*^VAR/VAR)^, respectively. All reprogramming proceeded as normal with first undifferentiated colonies appearing 7 days post transduction, stable colonies picked after 3 weeks and stable cell lines generated after approximately 40 days post transduction. Reprogrammed fibroblasts displayed the typical features of hiPSCs forming dense cell colonies, with well-defined colony boarders, containing cells with a high nuclear to cytoplasm ratio. To confirm their pluripotent status, Oct3/4 and NANOG expression was assessed as well as their ability to differentiate into the three embryonic germ layers using *in vitro* assays to differentiate cells into endoderm, mesoderm and ectoderm (Supplementary Figure 5D). Lastly, to ensure reprogramming had not caused any major chromosomal abnormalities we confirmed the karyotype as normal for both hiPSC lines (Supplementary Figure 5E and 5F).

### Wild-type and MTTP^(VAR/VAR)^ hiPSCs differentiate comparably but MTP levels are lower in the cell line expressing the variant protein and lipoprotein secretion is impaired

Next, to ensure our model would not be biased by different differentiation efficiency of our hiPSC lines, we differentiated cells from the study donors 1 and J into HLCs to compare morphology and gene expression profiles.^26^ Both cell lines appeared morphologically similar during all stages of differentiation and generated a monolayer of HLCs by day 21 (Figure 4A). Gene expression in both cell lines was similar at each of the developmental time points including definitive endoderm, foregut endoderm, and hepatoblast cells. Expression of genes associated with a mature hepatocyte phenotype (*HNF4α*, *ALB*, *A1AT*) was not significantly different between the two cell lines (Figure 4B). To further confirm HLCs from both hiPSCs were equivalent we performed mRNA-sequencing on day 21 cells and used EnrichR to query likely tissue and cell types based on gene expression from both populations of HLCs (Supplementary Table 4). For both HLC cell lines derived from the different donors, the top tissue hit was liver bulk tissue followed by hepatocyte. In terms of cell lines, both HLC cell lines were highly similar to hepatocyte cell lines HepG2, HUH7 and HEP3B. Importantly, while *MTTP* mRNA levels were not statistically different between *MTTP^(WT/WT)^* and *MTTP^(^*^VAR/VAR)^ HLCs (Figure 4B), immunocytochemistry showed high levels of MTP in those expressing the wild-type allele with only weak diffuse staining in *MTTP^(^*^VAR/VAR)^ HLCs suggesting reduced levels of the mutant protein (Figure 4C).

**Figure 4.**
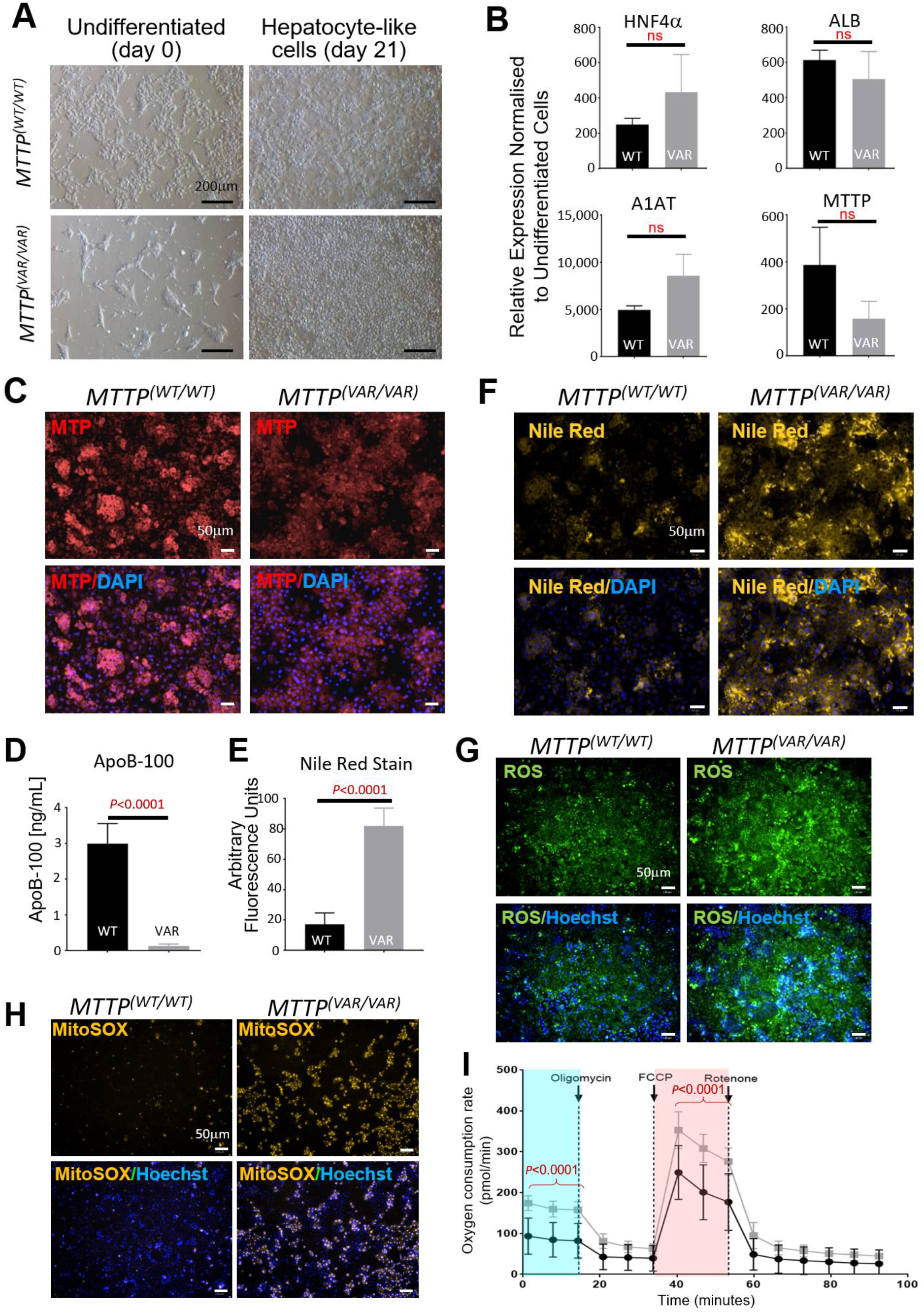
*MTTP*-564TT variant HLCs display oxidative stress and impaired mitochondrial metabolism. **(A)** Light microscopy showing terminally differentiated human induced pluripotent stem cell (hIPSC)-derived hepatocyte-like cells (HLCs) *MTTP^(WT/WT)^* (from MTP-564II healthy volunteer) and *MTTP*^VAR/VA*T)*^ (from MTP564-TT family member J). **(B)** Expression of mature hepatocyte markers: hepatocyte nuclear factor 4 alpha (HNF4α); albumin (ALB), alpha-1 antitrypsin (A1AT), and *MTTP* in *MTTP^(WT/WT)^* (black) and *MTTP^(^*^VAR/VAR)^ (grey) hiPSC derived HLCs quantified using RT-qPCR. **(C)** MTP protein expression revealed by immunocytochemistry ± DAPI staining. **(D)** Quantification of Nile Red fluorescence in derived *MTTP^(WT/WT)^* and *MTTP^(^*^VAR/VAR)^ cells. **(E)** ApolipoproteinB-100 (ApoB-100) quantification in cells by ELISA. **(F)** Nile Red staining of lipids using fluorescence microscopy ± DAPI stain. **(G)** Reactive oxygen species detected in cells by fluorescence microscopy ± Hoechst stain. **(H)** Superoxide presence in cells detected by fluorescence microscopy ± Hoechst stain. **(I)** Basal and maximal mitochondrial respiratory rates in *MTTP^(WT/WT)^* (●) and *MTTP^(^*^VAR/VAR)^ (□) hiPSC derived hepatocytes. The p values are comparisons between the cell lines at the time points indicated.

To assess VLDL export capabilities of the cell lines, and thus functioning of MTP in lipoprotein biosynthesis, levels of the hepatic protein constituent of VLDL, ApoB-100, were determined in the HLC culture medium after 48h of growth. Apo-B100 was detected in tissue culture medium from *MTTP^(WT/WT)^* HLCs as expected^36^, however there was significantly less ApoB-100 in the media from *MTTP^(VAR/VAR)^* HLCs confirming the clinical phenotype (Figure 4D) and that the MTP I564T variant in these patients affects ApoB-100 processing and lipid trafficking.

### MTTP^(VAR/VAR)^ HLCs inherently accumulate intracellular lipids

We cultured the derived HLCs to observe any phenotypic differences between the 2 cell lines. After growth for 48h in hepatocyte culture medium, *MTTP^(WT/WT)^* cells maintained a normal hepatocyte morphology and they accumulated a few apparent lipid droplets in the cytoplasmic space. In contrast, the *MTTP^(^*^VAR/VAR)^ HLCs appeared to develop discrete lipid droplets and clusters of many very small lipid droplets throughout the cytoplasm. To confirm these were lipid droplets we used the lipophilic dyes Oil-Red-O (Supplementary Figure 7C) and Nile Red to stain lipid vesicles demonstrating differences in the amount of accumulated lipid. Quantification of Nile Red fluorescence intensity showed levels were more than 4-fold higher in *MTTP^(^*^VAR/VAR)^ HLCs, (Figure 4E and 4F). This is consistent with the proposed reduced VLDL secretion in cells expressing MTP564-TT, restricting removal of intracellular triglycerides.

### Increased generation of ROS and altered mitochondrial respiration in MTTP^(VAR/VAR)^ HLCs

Hepatic free fatty acids can be converted to triglyceride for storage as cytoplasmic droplets or secreted as VLDL, or else directly metabolised via mitochondrial β-oxidation. Therefore, impaired MTP functionality restricting lipid secretion, thus increasing the availability of fatty acids, may impact on mitochondrial activities. Of importance, increased β-oxidation would generate additional ROS which can be a major driver of oxidative stress and cellular dysfunction. Cytoplasmic ROS were detected in both *MTTP^(WT/WT)^* and *MTTP^(^*^VAR/VAR)^ HLCs with levels being significantly higher in the *MTTP^(^*^VAR/VAR)^ cell line (Figures 4G, 5A and Supplementary Figure 7). Mitochondrial superoxide production was also significantly higher in *MTTP^(^*^VAR/VAR)^ HLCs (Figures 4H and 5B) consistent with increased fatty acid metabolism. This was further evidenced using mitochondrial stress testing measuring the oxygen consumption rate in live cells which found that *MTTP^(^*^VAR/VAR)^ HLCs had significantly higher basal and maximal mitochondrial respiration compared to mitochondria from the wild type cell line (Figure 4I).

**Figure 5.**
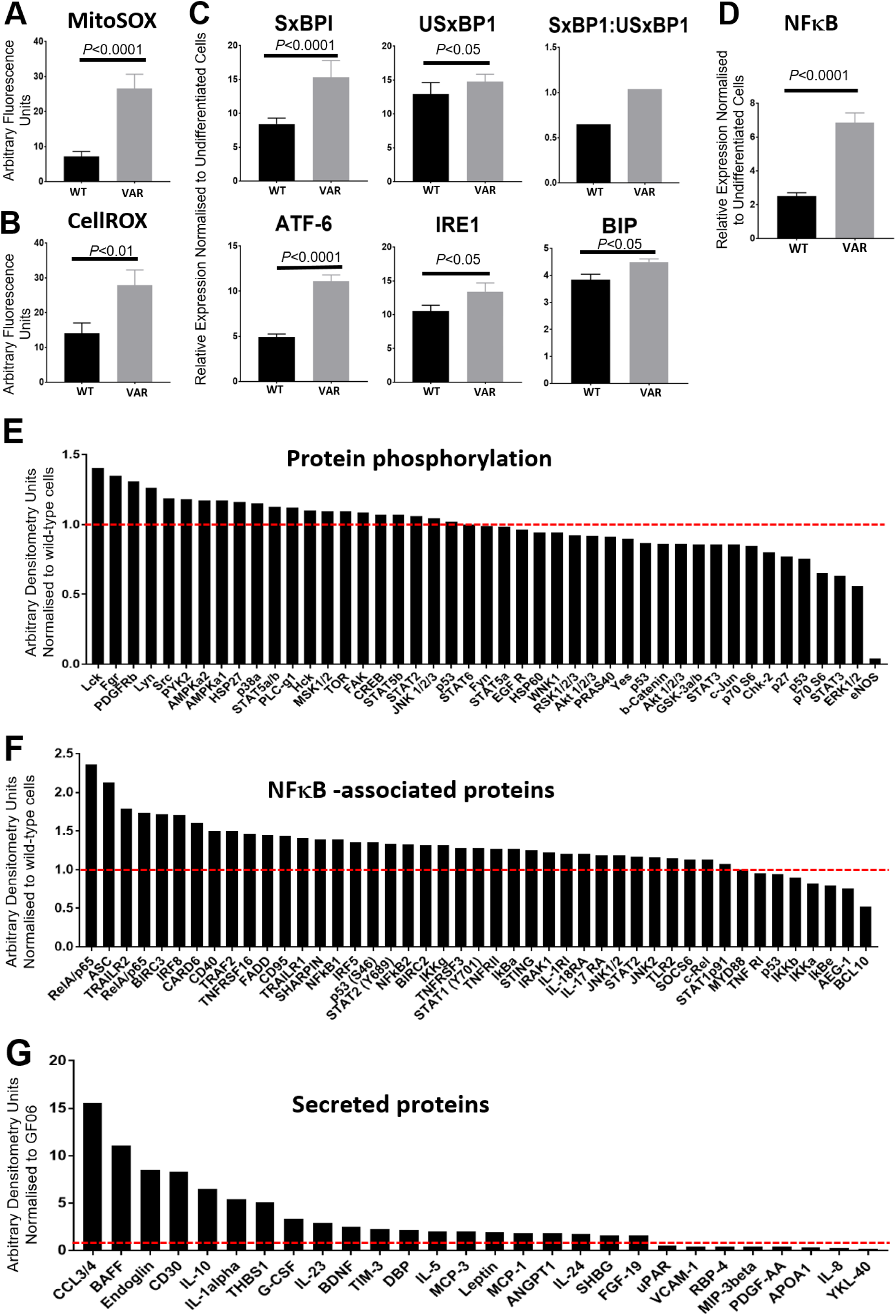
Phenotypic characterisation of *MTTP^(VAR/VAR)^* hiPSC derived HLCs. **(A)** Elevated oxidative stress indicated by comparison of MitoSOX fluorescence from *MTTP^(WT/WT)^* and *MTTP^(VAR/VAR)^* hepatocyte-like cells (HLCs). **(B)** Elevated mitochondrial stress indicated by comparison of CellROX fluorescence from *MTTP(WT/WT)* and *MTTP(VAR/VAR)* HLCs. **(C)** Elevated expression of ER-stress related genes determined by qPCR in *MTTP^(VAR/VAR)^* human induced pluripotent stem cell (hIPSC)-derived HLCs. **(D)** *NFκB* mRNA expression determined by qPCR. **(E)** Changes in protein phosphorylation in *MTTP^(VAR/VAR)^* hiPSC-derived HLCs. Expression normalised to *MTTP^(WT/WT)^* hiPSC derived HLCs quantified using densitometry of phosphokinase array. **(F)** Expression of NFκB-associated intracellular signalling proteins in *MTTP^(VAR/VAR)^* hiPSC derived HLCs, relative to expression in *MTTP^(WT/WT)^* HLCs. Expression quantified using densitometry of NFκB protein array. **(G)** Levels of proteins secreted into tissue culture medium in *MTTP^(VAR/VAR)^* hiPSC derived HLCs, normalised to *MTTP^(WT/WT)^* hiPSC derived HLCs quantified using densitometry of cytokine array.

### Increased NFκB signalling, inflammation, ER stress and secretion of pro-inflammatory mediators in MTTP^(VAR/VAR)^ HLCs

Impaired lipid trafficking and lipoprotein assembly incurred as a consequence of reduced MTP functionality is likely to cause a range of cellular responses including ER stress and inflammation. Steatosis is specifically associated with hepatic inflammation including activation of NFκB intracellular signalling pathways and secretion of pro-inflammatory mediators.^37^

Analysis of mRNA revealed that expression of ER stress mediators: spliced X-Box Binding protein-1 (SxBP1); activating transcription factor 6 (ATF6); binding immunoglobulin protein (BIP); and the ER stress transducer, inositol requiring enzyme 1 (IRE1) were significantly higher in *MTTP^(^*^VAR/VAR)^ HLCs (Figure 6C). In addition, *MTTP^(^*^VAR/VAR)^ HLCs expressed higher levels of NFκB (Figure 6D), Toll-like receptors TLR-3, 4 and 5, and TGFβ (data not shown), suggesting the cell line had an activated ER stress response and had active pro-inflammatory response compared to the wild-type cell line.

**Figure 6.**
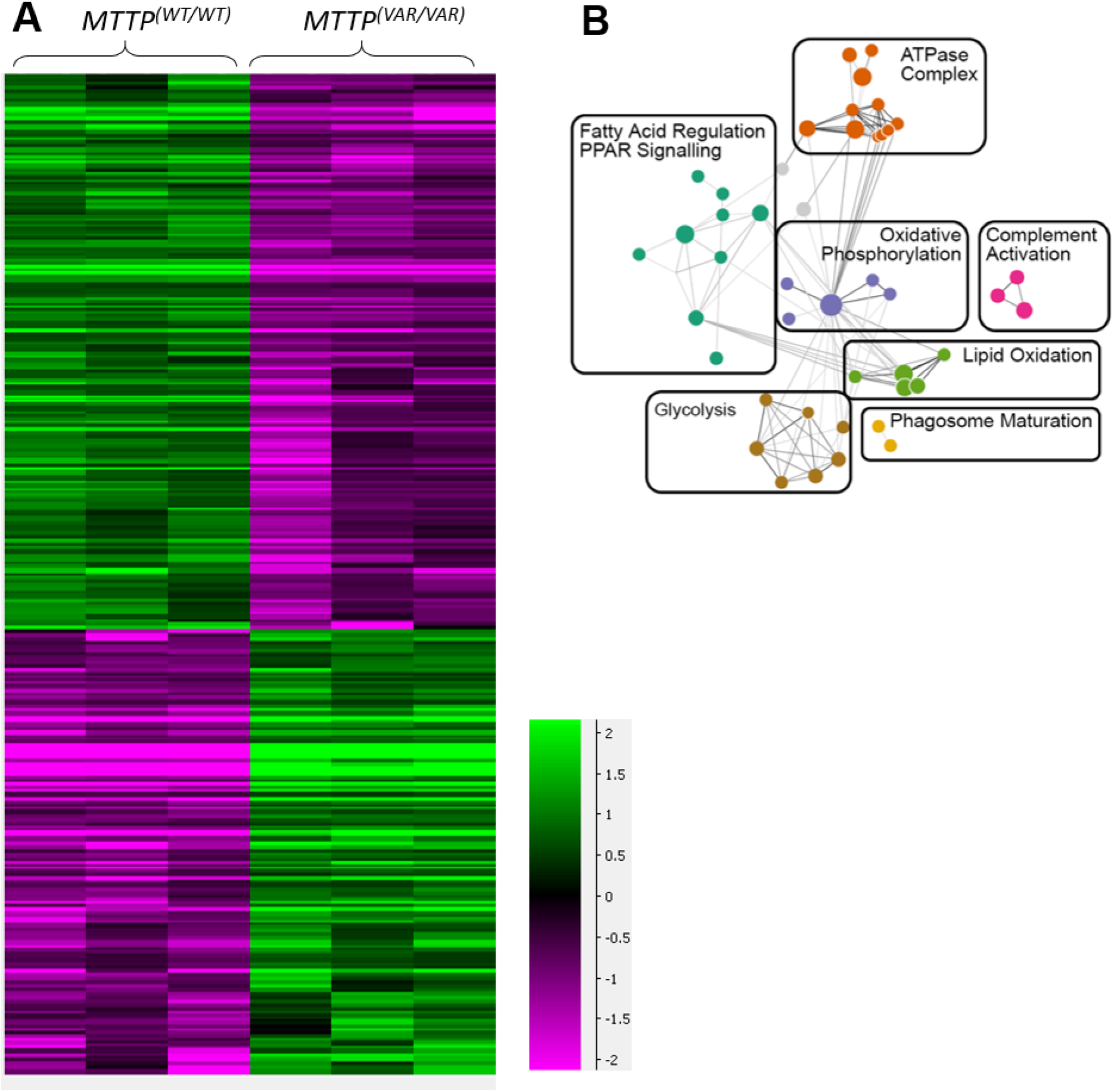
Comparison of expression patterns in HLCs with MTP564-II and MTP564-TT. **(A)** Heatmap representing differentially expressed genes in wild type and MTTP variant human induced pluripotent stem cells (hIPSCs) (in triplicate). **(B)** Giraph plot from quantitative gene set analysis showing gene sets up-regulated in *MTTP^(^*^VAR/VAR)^ hIPSC-derived hepatocyte-like cells (HLCs).

These observations were confirmed by assessing changes in intracellular and extracellular signalling using protein arrays to assess NFκB pathway activation, pro-inflammatory pathways, protein secretion and general protein phosphorylation (Figures 5, 6E, 6F and 6G). The observed lipid accumulation in the novel MTP564-TT variant strain was associated with widespread increases in NFκB pathway activation via phosphorylation of pro-inflammatory and pro-apoptotic pathway mediators including RelA/p65 complex, ASC, p53, FADD, and CD95. This increased pro-inflammatory signalling coincided with increased secretion of pro-inflammatory mediators including CCL3/4, BAFF, CD30, IL-1, IL-10 and IL-23, DBP and leptin while showing decreased RBP4 and PDGF. Notably, *MTTP^(^*^VAR/VAR)^ HLCs had more than 10-fold lower expression of phosphorylated-eNOS compared to *MTTP^(WT/WT)^* HLCs (Figure 6E). There were no significant differences in protein phosphorylation in the other 44 proteins assayed, between the 2 derived cell lines; however, there was a trend towards decreased ERK1/2 phosphorylation in *MTTP^(^*^VAR/VAR)^ HLCs.

### MTTP^(VAR/VAR)^ HLCs show increased expression of ECM remodelling and lipid metabolising genes

Lastly, to better understand genome wide changes in gene expression in *MTTP^(^*^VAR/VAR)^ and *MTTP^(WT/WT)^* HLCs we performed mRNA-sequencing following 2 days of culture and spontaneous lipid accumulation. Bioinformatics revealed 472 genes differentially expressed between the two HLC populations (Figure 6A) involved in diverse cellular functions including glycolysis, lipid oxidation, oxidative phosphorylation and complement activation and supporting previous observations of increased ROS generation and altered mitochondrial activity (Figure 4). Interestingly, Gene Ontology (GO) terms associated with the changing genes (Figure 6B) were mostly implicated in ECM remodelling, ECM organisation and degradation, ECM receptor interactions, and proteoglycan modifications, suggesting that ECM remodelling and fibrosis are very early features of hepatosteatosis and may precede other more well characterised markers of the disease (Supplementary Tables 5-7).

## Discussion

We have identified and characterised a rare *MTTP* variant (p.I564T) associated with an inherited form of non-alcoholic fatty liver disease, in a four-generation family. Our investigation has revealed a variant resulting in decreased ApoB-containing lipoprotein secretion in homozygotes (demonstrated in persons E, F and J of the family studied), rather than abetalipoproteinemia where ApoB is undetectable (Figure 2B, Table 1)^7, 9, 38–40^. Levels of ApoB in heterozygotes K, Q and a further five heterozygotes in generation II were normal. Although other carriages of this variant have been described, no phenotypic characteristics related to these are previously reported^29^, gnomAD). Protein modelling (Figure 1B; Ensembl) suggests the substitution affects the protein structure but does not lie in the active site or in functional regions previously described.^22, 41, 42^ The T-C change possibly impacts on mRNA or protein stability/turnover or alter partner interactions which would reduce or alter its activity resulting in a different phenotype to that of abetalipoproteinemia. Presentation of homozygote cases is clearly distinct from ABL^8, 38^ supporting suggested subtle phenotype whereby impact is limited to liver lipid imbalance. This provides potential for treatment through reduced dietary fat intake and makes it an attractive model for cellular consequences of lipid accumulation.

Phenotyping studies demonstrated contrasting levels of fasting serum ApoB levels as well as VLDL secretion following meal challenge in the two MTTP-564TT family members; while both of the biomarkers were substantially low in untreated individual F, these were in the normal range in the liver transplant recipient J (Figure 2B; Table 1). Previously, the *MTTP* -493 variant (rs1800591) G allele associated with reduced MTP function has been associated with NAFLD susceptibility in a meta-analysis of 11 case-control studies.^6^ Also an association study in non-diabetic patients with NASH (and controls) found GG homozygotes had significantly higher plasma triglycerides, intestinal and hepatic large VLDL and oxidised LDL than GG/GT group.^43^ Additionally, a candidate gene study found that in healthy men with elevated triglyceride levels, the MTTP p.H297Q variant was associated with higher ApoB levels.^44^

All of the five MTP564-IT heterozygous individuals showed ApoB and lipoprotein levels within the normal range. This is consistent with previous report that a single copy of MTTP may be sufficient for normal functionality.^45^ Disruption of ApoB biosynthesis and associated VLDL secretion has been widely described with a spectrum of consequences linked to characterised pathologies.^10^ The underlying mechanisms are inherently linked to nutritional intake with diets high in fats (increasing hepatic fat content) and carbohydrate (increasing hepatic de novo lipogenesis) resulting in hyperlipidemia. Hepatic lipid balance is dependent on secretion of VLDL which is restricted by availability/activity of MTP, so any variants with altered activities are likely to have metabolic effects. The small number of people carrying the rare variant available for analysis is a limitation of this work.

Overall, VLDL secretion may increase with hepatic steatosis related to metabolic syndrome.^46^ However, in people carrying the PNPLA3 rs738409 G allele, a relatively decreased VLDL secretion per amount of liver fat (particularly triglyceride-rich, large VLDL) has been reported in non-diabetics^35^ and PNPLA3-mediated lipid remodelling linked to NAFLD development through unknown mechanisms.^47, 48^ VLDL secretion is also lowered in *TM6SF2* T carriers^33, 34, 49^. In our study, postprandial VLDL secretion, the predominant postprandial lipoprotein associated with hyperlipidemia^50^, was found to be reduced in cases homozygous for *TM6SF2* or *PNPLA3* but fasting ApoB levels were normal in contrast with *MTTP* p.I564T homozygosity where we observed both a reduced level of ApoB and VLDL-associated lipids.

In addition to demonstrating the functional consequences of *MTTP* p.I564T variant, the HLCs derived from hiPSCs provide a disease model for early stage NAFLD, beyond this triglyceride accumulation. However, we acknowledge that this model is derived from a single homozygote and compared to cells from an unrelated wild type donor. Studies have shown a link between the amount of steatosis, fibrosis development and liver disease mortality^51^ with lipid metabolism acting as the initiator of progression to NASH.^52^ Although sequestered triglycerides are suggested to provide a protective buffer, lipotoxicity may arise from metabolites such as saturated fatty acids leading to pathway components (including ACC-1/2, FXR/FGF19/FXR4, and SCD-1) being tested as therapeutic targets.^53^ Increased mitochondrial fatty acid β-oxidation may provide a protective response but uncontrolled results in the generation of ROS which can be a major driver of oxidative stress and cellular dysfunction (Figure 4I, Figure 6).

We show that as lipid accumulation increases, hepatocytes have increased ER stress, activate pro-inflammatory signalling pathways including NFκB, P53, eNOS and secrete pro-inflammatory mediators. This coincides with increased production of reactive oxygen species, superoxide production and alterations to mitochondrial respiration driving the disease progression leading to cirrhosis and hepatocellular carcinoma as seen among the family members. Similar findings were reported in cardiomyocytes derived in an MTTP pR46G variant model.^36^ Excessive lipid accumulation in hepatocytes can serve as substrates for the generation of lipotoxic species. One of the major consequences of hepatic lipid metabolism is mitochondrial β-oxidation and esterification to form triglycerides which can serve as a protective mechanism against lipotoxicity in hepatocytes. However, if lipid accumulation is in excess of the β-oxidation capacity, such as in NAFLD, toxic intermediates can accumulate which induce metabolic stress and subsequent inflammation and cell death.

We conclude that the main feature of the *MTTP* p.I564T variant is impaired ApoB secretion and hepatic lipid accumulation distinct from classical abetalipoproteinemia phenotype where MTP expression is abolished. Identification and characterization of rare disease such as hereditary NAFLD is of medical significance in Indian population where high rates of founder events have been reported.^54^ In addition, HLC modelling supports this providing additional details of signalling, inflammatory and metabolic cellular pathways involved, highlighting pathophysiology driving NAFLD progression and possible therapeutic targets.

## Supporting information

research reporting checklist

## Data Availability

The data that support the findings of this study are available on request from the corresponding author. Exome sequence data will be available for restricted access via The European Genome-phenome Archive.

## Acknowledgements

The views expressed are those of the authors and not necessarily those of the National Health Service (NHS), the NIHR or the Department of Health. We thank all the research participants particularly the family involved. We are grateful to the study teams of the EXCEED study, the Trivandrum cohort and NASH study for their contributions. We are grateful to the clinical team at University Hospitals Leicester NHS Trust for clinical work-up and acknowledge support from the late Roger Williams. We thank the high-throughput genomics group at the Wellcome Trust Centre for Human Genetics for the generation of the sequence data. We thank Michael Steward for generating illustration of MTP, and thank Sally Cordon and Ian Macdonald for assistance with metabolic analysis and Melanie Lingaya and Calum Greenhalgh for technical support. We thank Ester Burden-Teh and Jane Chalmers for assistance with taking skin biopsies and Antonella Ghezzi for obtaining clinical samples for genotyping. We thank Beth Robinson and Nottingham Digestive Diseases Team for assistance with coordinating participant involvement and the meal study. For the purpose of open access, the author has applied a CC BY public copyright licence to any Author Accepted Manuscript version arising from this submission.

## Sources of Funding

This work was supported by the Medical Research Council Nottingham Molecular Pathology Node [grant number MR/N005953/1], National Institute of Health Research Nottingham Digestive Diseases Biomedical Research Unit and Nottingham Biomedical Research Centre [BRC-1215-20003]. All cell modelling was supported by the RoseTrees Trust and the Stoneygate Trust [M546]. L. Wain holds a GSK/British Lung Foundation Chair in Respiratory Research (C17-1). The research was supported by the NIHR Leicester Biomedical Research Centre; C. John holds a Medical Research Council Clinical Research Training Fellowship [MR/P00167X/1]. The exome sequencing was funded by MRC Grant Senior Clinical Fellowship to M. Tobin (G0902313) and we thank the high-throughput genomics group at the Wellcome Trust Centre for Human Genetics (funded by Wellcome Trust Grant 090532/Z/09/Z and MRC Hub Grant G090074791070) for the generation of the sequence data. M. Tobin is supported by a Wellcome Trust Investigator Award [WT202849/Z/16/Z] and holds an NIHR Senior Investigator Award. The funders had no role in study design, data collection and analysis, decision to publish, or preparation of the manuscript.

## Notes

**Conflicts of Interest** Competing interests: All authors have completed the ICMJE uniform disclosure form at www.icmje.org/coi_disclosure.pdf and declare: no support from any organization for the submitted work; Guruprasad Aithal has received unrelated research grants from Pfizer Inc, and served as a consultant and an advisory board member for Pfizer Inc, GlaxoSmithKline and Median Technologies and is President of the British Association for the Study of the Liver; Louise Wain has received unrelated research grants from GlaxoSmithKline and Orion Pharma Ltd; Martin Tobin has unrelated research collaborations with GlaxoSmithKline and Orion Pharma Ltd; Ioanna Ntalla is employed by Gilead Sciences Ltd. (since August 2019). no other relationships or activities that could appear to have influenced the submitted work.

### Competing Interest Statement

Competing interests: All authors have completed the ICMJE uniform disclosure form at www.icmje.org/coi_disclosure.pdf and declare: no support from any organization for the submitted work; Guruprasad Aithal has received unrelated research grants from Pfizer Inc, and served as a consultant and an advisory board member for Pfizer Inc, GlaxoSmithKline and Median Technologies and is President of the British Association for the Study of the Liver; Louise Wain has received unrelated research grants from GlaxoSmithKline and Orion Pharma Ltd; Martin Tobin has unrelated research collaborations with GlaxoSmithKline and Orion Pharma Ltd; Ioanna Ntalla is employed by Gilead Sciences Ltd. (since August 2019). no other relationships or activities that could appear to have influenced the submitted work.

### Author Declarations

The meal-response study was approved by the Health Research Authority after review by National Research Ethics Service Committee North-East (Ref 16/NE/0251) and was sponsored by Nottingham University Hospitals NHS Trust. The Genetics of Rare Inherited Disorders (GRID) study was approved by the Health Research Authority after review by National Research Ethics Service East Midlands Northampton Committee (Ref 12/EM/0262).

