## supplemental material for "Identification and functional characterisation of a rare *MTTP* variant underlying hereditary non-alcoholic fatty liver disease"

Grove, J.I. et al.

#### Supporting Information

##### Supplemental Methods

**Table of Primers Used:**

| Variant | Primer Pair |
|---|---|
| rs745447480 | *MTTP*-F1: 5′- TCTTAACGGCCTCAGCCTAG<br>*MTTP*-R1: 5′- CAGAGTTACCAGTCATGGACTC |
| rs1800591 | *MTTP*-F2: 5′- GGCTTGCTAGTGTGCTAATGACAG<br>*MTTP*-R2: 5′- GAGTGACCCTCTTCAGAACCTGC |
| rs2306986 and rs3816873 | *MTTP*-F5: 5′- AAGGTAGAATAGGGCAGGGGTCC<br>*MTTP*-R5: 5′- CTAATCTCAGTTGGATCATTTCAGTCTC |
| rs3792683 | *MTTP*-F6: 5′- GTTACAGGTAGAGAACATGCTGACATG<br>*MTTP*-R6: 5′- CCTCCATGGTACAGTGGTGCAC |
| rs2306985 | *MTTP*-F7: 5′- CAGTCACAGAGTCCTACCCAGG<br>*MTTP*-R7: 5′- GAGACTGCTGTCATCACAACTCTGTG |
| rs738409 | PNPLA3-6R: 5′- CAGCTGTGGCTACTCTGTCTG<br>PNPLA3-4F: 5′-TGGAGAAAGCTTATGAAGGATCAG |
| rs1260326 | GCKR-2F: 5′- GGGTCTTAGGGTACCTGCTCAGAGG<br>GCKR-2R: 5′- GGTAACCCATGACCTTGCCCAGC |
| rs58542926 | TM6SF2-2F: 5′- CCAAAATGTTGGGATTACAGG<br>TM6SF2-2R: 5′- ACAGATGTCCAGCAGGGTTC |

**Genotyping:** For analysis of *MTTP* alleles in family members, the sequence of PCR products generated using the primers listed was determined using Sanger sequencing. PCR-RFLP genotyping of Indian cohort for rs745447480 used primers *MTTP*-F1 and *MTTP*-R1 followed by restriction digestion with Hpy166II. PCR-RFLP was used for genotyping rs738409 (FokI), rs58542926 (MspI) and rs58542926 (Hpy188I).

**Table of Antibodies Used:**

| Antibody | Source |
|---|---|
| MTTP | Abcam |
| OCT3/4 | Santa Cruz Biotechnology |
| NANOG | R&D Systems |
| MESP1 | Abcam |
| GATA-4 | R&D Systems |
| Nestin | Merck |
| SOX2 | Novus Biologicals |
| ALB | R&D Systems |
| CYP2A6 | OriGene Technologies |
| A1AT | Abcam |
| GST-pi | Enzo Life Sciences |
| Donkey Anti-Mouse AF 488 (1/400 dilution) | Invitrogen |
| Donkey Anti-Rabbit AF 647 (1/400 dilution) | Invitrogen |
| Donkey Anti-Goat AF 647 (1/400 dilution) | Invitrogen |

**Microscopy:** Cells were fixed in 4% paraformaldehyde (PFA; VMR) for 20 min at 4°C. Fixed cells were stained with 0.5 µg/ml DAPI (Sigma), 5 µg/ml Hoechst 33342 (Invitogen) in 1% FBS-PBST for 5 min to counter-stain nuclei, or 30µM Nile Red in methanol (Invitrogen) for 15min in the dark or Oil-Red-O. For immunocytochemistry, blocking and membrane permeabilization of fixed cells was done using 10% foetal bovine serum (FBS; Gibco) and 0.1% TritonX-100 (Thermo Scientific) in PBS (Gibco) for 30min. Cells were incubated overnight at 4°C in a combination of primary antibodies diluted in 1% FBS-PBST solution. The next day, cells were incubated with fluorescent-labelled secondary antibodies for 1h at ambient temperature, followed by nuclei-counterstaining using 0.5 µg/ml DAPI (Sigma) in 1% FBS-PBST for 5min. Images were acquired using the Operetta high content image analyser and analysed with Columbus system (both PerkinElmer).

### Supplemental Tables

**Supplemental Table 1.** Clinical characteristics of other individuals investigated.

| MTTP genotype[1]: MTP residue 564 (variant=T) | Sex | Liver Disease Diagnosis (method) | T2D | BMI | Hypertension | Liver biochemistry (LFTs) & lipid blood analyses at diagnosis |
|---|---|---|---|---|---|---|
| **I T** | M | NASH (USS) | x | <25 | x | Elevated ALT, Elevated lipids<br>TE<7.9kPa CAP>302 |
| **I T** | M | FL (USS) | x | <25 | x | Normal LFTs, Normal Lipids; |
| **I T** | M | FL (USS) | x | <30 | x | Elevated ALT, Elevated lipids<br>TE<7.9kPa CAP>302 |
| **I T** | F | healthy (USS) | x | <25 | x | Normal LFTs, Normal Lipids<br>TE<7.9kPa CAP<302 |
| **I T** | M | healthy (TE) | x | <30 | x | Normal Lipids; TE<7.9kPa |
| **I T** | M | healthy (USS) | x | <25 | x | Normal LFTs, Normal lipids, |
| **I T** | F | healthy (USS) | x | <25 | x | Normal LFTs, Normal lipids,<br>TE<7.9kPa CAP<302 |
| **I T** | F | healthy (USS) | x | <25 | x | Normal LFTs, Normal Lipids; |
| **I T**[2] | F | healthy (USS) | x | <25 | x | Normal LFTs, Normal lipids,<br>TE<7.9kPa CAP<302 |
| **I T**[2] | M | healthy (USS) | x | <30 | x | Normal LFTs, Normal lipids,<br>TE<7.9kPa CAP<302 |
| **I T** | M | healthy (USS) | x | <30 | x | Normal LFTs (ALT=14), Normal lipids,<br>TE<7.9kPa CAP<302 |
| **I T** | F | FL (USS) | x | <25 | x | Normal LFTs, Elevated lipids, TE<7.9kPa CAP>302 |
| | | healthy (USS) | | | | Recovered. TE<7.9kPa CAP<302 |
| **I T** | M | FL (USS) | x | <30 | x | Normal LFTs, Normal Lipids |
| | | healthy (USS) | | | | Recovered.<br>TE<7.9kPa CAP<302 |
| **I I**[3] | M | FL (USS) | x | <25 | x | Normal LFTs, Normal Lipids |
| | | healthy (USS) | | | | Recovered. TE<7.9kPa CAP<302 |
| **I I**[3] | F | healthy (USS) | x | <25 | x | Normal LFTs, Normal lipids,<br>TE<7.9kPa CAP<302 |
| **I I**[3] | F | healthy (USS) | x | <30 | x | Normal LFTs, Normal Lipids |

[1] *MTTP* I564T novel variant (mutant allele nucleotide C results in threonine residue 564 (T)); [2] likely heterozygote as parent was MTP564-TT. F: female; M: male; USS: abdominal ultrasonographic steatosis score; NASH: non-alcoholic steatohepatitis; FL: fatty liver; T2D: Type 2 diabetes mellitus; LFTs: liver function tests; ALT: alanine transaminase U/L; TE: transient elastography; CAP: Controlled Attenuation Parameter dB/m. Further clinical details are available upon request to the corresponding author.

**Supplemental Table 2.** *MTTP* genotyping

| Family member | MTTP p.I564T rs745447480 (T>C) | PNPLA3 p.I148M rs738409 (C>G) | TM6SF2 p.E167K rs58542926 (C>T) | MTTP -493 rs1800592 (G>T) | MTTP p.I128T rs3816873 (T>C) | MTTP Q297H rs2306985 (G>C) |
|---|---|---|---|---|---|---|
| B ♀ | I T (TC) | M M (GG) | E K (CT) | TT | T T (CC) | H H (CC) |
| C ♀ | T T (CC) | I M (CG) | E E (CC) | TT | T T (CC) | H H (CC) |
| D ♂ | T T (CC) | I M (CG) | E E (CC) | TT | T T (CC) | H H (CC) |
| E ♂ | I T (TC) | I M (CG) | E E (CC) | TT | X | H H (CC) |
| F ♂ | T T (CC) | I M (CG) | E K (CT) | TT | T T (CC) | H H (CC) |
| G ♂ | X | X | X | X | X | X |
| H ♂ | T T (CC) | I M (CG) | E K (CT) | TT | T T (CC) | X |
| I ♀ | I I (TT) | I M (CG) | E E (CC) | TG | T T (CC) | H H (CC) |
| J ♀ - | T T (CC) | I M (CG) | E E (CC) | TT | T T (CC) | H H (CC) |
| K ♀ | I T (TC) | I M (CG) | E K (CT) | TG | X | H H (CC) |
| L ♀ | T T (CC) | I M (CG) | E E (CC) | TT | T T (CC) | H H (CC) |
| M ♂ | I I (TT) | I I (CC) | E K (CT) | TG | I T (TC) | Q H (GC) |
| N ♂ | I T (TC) | I M (CG) | E K (CT) | TG | X | Q H (GC) |
| O ♂ | I T (TC) | I I (CC) | E K (CT) | TT | I T (TC) | Q H (GC) |
| Q ♂ | I T (TC) | M M (GG) | E E (CC) | TG | I T (TC) | H H (CC) |

Family pedigree showing relationship between family members is available on request from the corresponding author. The genotype allele is shown in parentheses; light grey indicates heterozygote; dark grey indicates homozygous for effect allele. Blue letters are amino acids encoded. For E98D rs2306986 (C): all tested were GG; For N166S rs3792683 (G): all tested were AA. X = not determined.

**Supplemental Table 3.** Characteristics of study participants included in meal response analysis.

| **Family member studied** | | **Matched healthy volunteer** | | **Matched NAFLD patient** | |
|---|---|---|---|---|---|
| **Person ID-Family** | Description | **Person ID-Healthy Volunteer** | Description | **Person ID-NAFLD case** | Description |
| **F** | ♂ BMI <25, **MTP564-TT** cirrhosis, PNPLA3-IM TM6SF2-EK | **1** | ♂ BMI <30, PNPLA3-IM TM6SF2-EE | **2** | ♂ BMI ≥30, NASH + fibrosis PNPLA3-IM TM6SF2-EK |
| | | | | **3** | ♂ BMI <30, NASH + fibrosis PNPLA3-IM **TM6SF2-KK** |
| **J** | ♀ BMI <25, **MTP564-TT** healthy transplanted PNPLA3-IM TM6SF2-EE | **4** | ♀ BMI <30, **PNPLA3-MM** TM6SF2-EE | **5** | ♀ BMI <30, NASH no fibrosis, PNPLA3-II TM6SF2-EE |
| **K** | ♀ BMI <25, MTP564-IT fatty liver PNPLA3-IM TM6SF2-EK | **6** | ♀ BMI <30, PNPLA3-II TM6SF2-EE | **7** | ♀ BMI ≥30, NASH + fibrosis, **PNPLA3-MM** TM6SF2-EE |
| **M** | ♂ BMI <30, MTP564-II healthy PNPLA3-II TM6SF2-EK | **8** | ♂ BMI <30, PNPLA3 CC TM6SF2 CT | **9** | ♂ BMI ≥30, NASH + fibrosis, diabetic PNPLA3-II TM6SF2-EE |
| **Q** | ♂ BMI <25, MTP564-IT healthy **PNPLA3-MM** TM6SF2-EE | **10** | ♂ BMI <30, PNPLA3-II TM6SF2-EE | **11** | ♂ BMI ≥30, NASH + fibrosis **PNPLA3-MM** TM6SF2-EK |

Participants were gender matched to family members and age matched within 9 years. Patients had NAFLD diagnosis on liver biopsy. Skin biopsy samples from participants J and 1 were used to derive cell lines used for analysis. *MTTP* mutation C encodes MTP-564T. *PNPLA3* C>G rs738408 and *TM6SF2* C>T rs58542926 genotype is shown. *PNPLA3* G is the effect allele encoding 148M; *TM6SF2* T is the effect allele encoding 167K. Bold indicates homozygous effect alleles.

**Supplemental Table 4.** All RNA-seq and CHIP-seq Sample Search Space (ARCHS4) tissue type and cell type predictions for hIPSC-derived hepatocyte-like cells.

| Index | | MTP564-II wild type hIPSC derived hepatocyte-like cells | | | | | MTP564-TT variant hIPSC derived hepatocyte-like cells | | | | |
|---|---|---|---|---|---|---|---|---|---|---|---|
| | | Name | P-value | Adjusted P-value | Odds ratio | Combined Score | Name | P-value | Adjusted P-value | Odds ratio | Combined Score |
| Tissue Type Prediction | 1 | Liver (bulk tissue) | 6.662e-163 | 7.194e-161 | 2.72 | 1017.19 | Liver (bulk tissue) | 2.363e-171 | 2.552e-169 | 2.74 | 1075.07 |
| | 2 | Hepatocyte | 3.025e-118 | 1.633e-116 | 2.44 | 661.17 | Hepatocyte | 4.413e-123 | 2.383e-121 | 2.45 | 689.05 |
| | 3 | Small intestine (bulk tissue) | 1.268e-78 | 4.564e-77 | 2.15 | 385.83 | Small intestine (bulk tissue) | 4.644e-82 | 1.672e-80 | 2.16 | 403.60 |
| | 4 | Gastric epithelial cell | 9.568e-70 | 2.583e-68 | 2.08 | 330.09 | Ileum (bulk tissue) | 4.053e-68 | 1.094e-66 | 2.04 | 316.85 |
| | 5 | Ileum (bulk tissue) | 4.833e-67 | 1.044e-65 | 2.05 | 313.59 | Lung (bulk tissue) | 1.365e-61 | 2.948e-60 | 1.99 | 278.23 |
| | 6 | Lung (bulk tissue) | 5.792e-58 | 1.042e-56 | 1.97 | 259.88 | Gastric epithelial cell | 1.754e-59 | 3.158e-58 | 1.97 | 266.04 |
| | 7 | Colon (bulk tissue) | 1.893e-53 | 2.921e-52 | 1.93 | 234.18 | Colon (bulk tissue) | 2.340e-53 | 3.610e-52 | 1.91 | 231.44 |
| | 8 | Gastric tissue (bulk) | 4.762e-46 | 6.428e-45 | 1.85 | 193.58 | Gastric tissue (bulk) | 3.668e-47 | 4.952e-46 | 1.85 | 197.74 |
| | 9 | Omentum | 3.685e-39 | 4.422e-38 | 1.78 | 157.60 | Omentum | 6.260e-45 | 7.512e-44 | 1.83 | 185.93 |
| | 10 | Amniotic fluid | 1.454e-23 | 1.570e-22 | 1.59 | 83.40 | Skin (bulk tissue) | 5.090e-28 | 5.497e-27 | 1.63 | 102.71 |
| Cell Type Prediction | 1 | HEPG2 | 9.067e-83 | 1.133e-80 | 2.16 | 407.91 | HEPG2 | 1.419e-75 | 1.774e-73 | 2.08 | 358.54 |
| | 2 | HUH7 | 5.774e-52 | 3.609e-50 | 1.90 | 223.63 | HUH7 | 5.473e-42 | 3.421e-49 | 1.87 | 216.27 |
| | 3 | HEP3B | 1.702e-41 | 7.091e-40 | 1.79 | 168.03 | HEP3B | 9.101e-42 | 3.754e-40 | 1.78 | 167.99 |
| | 4 | CFPAC1 | 1.049e-38 | 3.278e-37 | 1.76 | 153.90 | CFPAC1 | 2.518e-38 | 7.870e-37 | 1.74 | 150.72 |
| | 5 | CAPAN1 | 5.244e-36 | 1.311e-34 | 1.73 | 140.51 | MCF10 | 2.035e-33 | 5.088e-32 | 1.69 | 126.93 |
| | 6 | A549 | 1.156e-29 | 2.409e-28 | 1.65 | 110.23 | CAPAN1 | 3.661e-32 | 7.626e-31 | 1.67 | 121.00 |
| | 7 | BXPC3 | 7.111e-28 | 1.270e-26 | 1.63 | 102.00 | A549 | 7.477e-32 | 1.335e-30 | 1.67 | 119.54 |
| | 8 | MCF10 | 2.724e-25 | 4.257e-24 | 1.60 | 90.38 | BXPC3 | 4.377e-27 | 6.840e-26 | 1.61 | 97.69 |
| | 9 | RT4 | 1.230e-23 | 1.709e-22 | 1.58 | 83.09 | RT4 | 1.444e-24 | 2.006e-23 | 1.58 | 86.55 |
| | 10 | HT29 | 2.676e-22 | 3.345e-21 | 1.56 | 77.31 | HNSCC | 1.740e-23 | 2.175e-22 | 1.56 | 81.86 |

**Supplemental Table 5.** Gene set analysis showing terms for all genes differentially expressed between MTP564-II wild type and MTP564-TT variant hIPSC-derived hepatocyte-like cells.

| source | term_name | term_id | adjusted_p_value | negative_log10_of_adjusted_p_value |
|---|---|---|---|---|
| REAC | Extracellular matrix organization | REAC:R-HSA-1474244 | 4.87E-07 | 6.312667258 |
| GO:CC | extracellular matrix | GO:0031012 | 2.85941E-06 | 5.543723748 |
| REAC | Degradation of the extracellular matrix | REAC:R-HSA-1474228 | 2.43072E-05 | 4.614264431 |
| GO:BP | extracellular matrix organization | GO:0030198 | 0.000831626 | 3.080071732 |
| GO:BP | extracellular structure organization | GO:0043062 | 0.000894227 | 3.048552438 |
| REAC | Collagen formation | REAC:R-HSA-1474290 | 0.003136295 | 2.503583115 |
| GO:BP | digestion | GO:0007586 | 0.004959299 | 2.304579689 |
| GO:CC | collagen-containing extracellular matrix | GO:0062023 | 0.00516165 | 2.287211438 |
| KEGG | Protein digestion and absorption | KEGG:04974 | 0.006978005 | 2.156268728 |
| REAC | Collagen biosynthesis and modifying enzymes | REAC:R-HSA-1650814 | 0.014030082 | 1.852939779 |
| KEGG | ECM-receptor interaction | KEGG:04512 | 0.014057765 | 1.852083733 |
| REAC | ECM proteoglycans | REAC:R-HSA-3000178 | 0.016110075 | 1.792902441 |
| TF | Factor: slug; motif: NRCAGGTGCR; match class: 1 | TF:M12259_1 | 0.027606305 | 1.558991718 |

**Supplemental Table 6.** Gene set analysis showing terms for genes upregulated in MTP564-II wild type hIPSC derived hepatocyte-like cells compared to MTP564-TT variant hIPSC-derived hepatocyte-like cells.

| source | term_name | term_id | adjusted_p_value | negative_log10_of_adjusted_p_value |
|---|---|---|---|---|
| GO:CC | midbody | GO:0030496 | 0.014234541 | 1.846656538 |
| REAC | Resolution of Sister Chromatid Cohesion | REAC:R-HSA-2500257 | 0.018667177 | 1.728921362 |
| REAC | Amplification of signal from unattached kinetochores via a MAD2 inhibitory signal | REAC:R-HSA-141444 | 0.023621746 | 1.626688011 |
| REAC | Amplification of signal from the kinetochores | REAC:R-HSA-141424 | 0.023621746 | 1.626688011 |
| GO:CC | spindle | GO:0005819 | 0.026656468 | 1.5741974 |

**Supplemental Table 7.** Gene set analysis showing terms for genes upregulated in MTP564-TT variant hIPSC derived hepatocyte-like cells compared to MTP564-II wild type hIPSC-derived hepatocyte-like cells.

| source | term_name | term_id | adjusted_p_value | negative_log10_of_adjusted_p_value |
|---|---|---|---|---|
| REAC | Extracellular matrix organization | REAC:R-HSA-1474244 | 0.000145038 | 3.838517822 |
| REAC | Degradation of the extracellular matrix | REAC:R-HSA-1474228 | 0.000353562 | 3.451534414 |
| REAC | ECM proteoglycans | REAC:R-HSA-3000178 | 0.002952027 | 2.529879617 |
| GO:BP | extracellular matrix organization | GO:0030198 | 0.008885339 | 2.051326005 |
| GO:BP | extracellular structure organization | GO:0043062 | 0.009318155 | 2.030670061 |
| HP | Abnormal cardiovascular system physiology | HP:0011025 | 0.014922899 | 1.826146793 |
| KEGG | ECM-receptor interaction | KEGG:04512 | 0.01921473 | 1.71636572 |
| KEGG | Focal adhesion | KEGG:04510 | 0.027077396 | 1.567393098 |
| WP | Focal Adhesion | WP:WP306 | 0.030961443 | 1.509178802 |
| HP | Osteoporosis | HP:0000939 | 0.038409054 | 1.415566389 |
| HP | Abnormality of skin physiology | HP:0011122 | 0.043372457 | 1.362785975 |
| HP | Abnormality of humoral immunity | HP:0005368 | 0.043717346 | 1.35934621 |
| HP | Abnormal vascular physiology | HP:0030163 | 0.04508747 | 1.345944131 |

#### Supplemental Figures

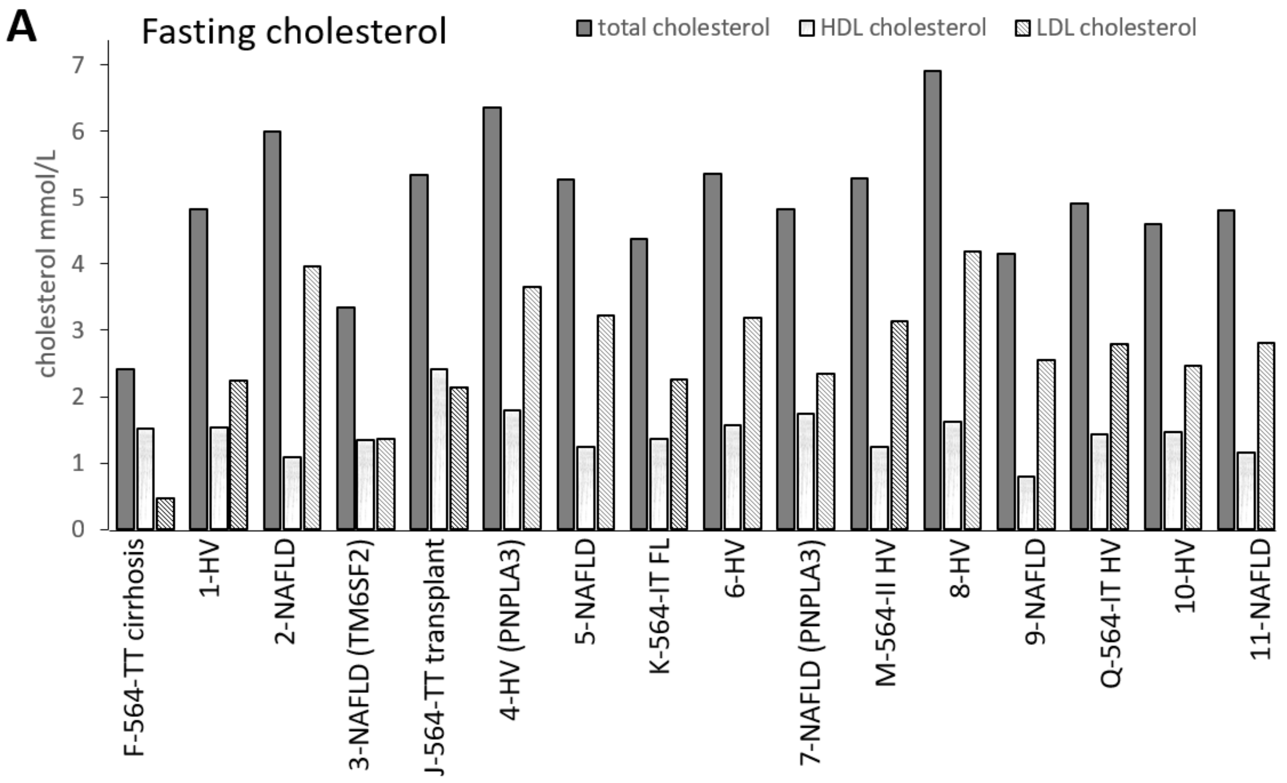


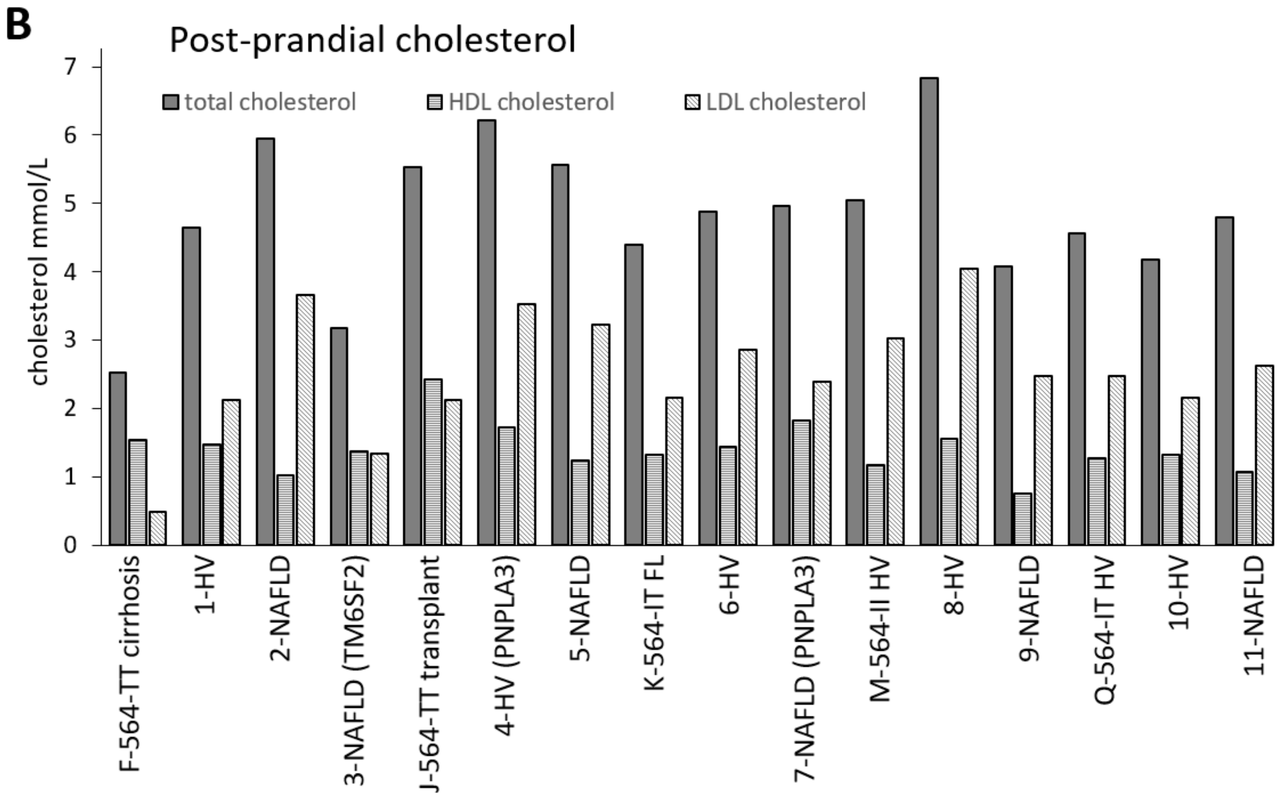

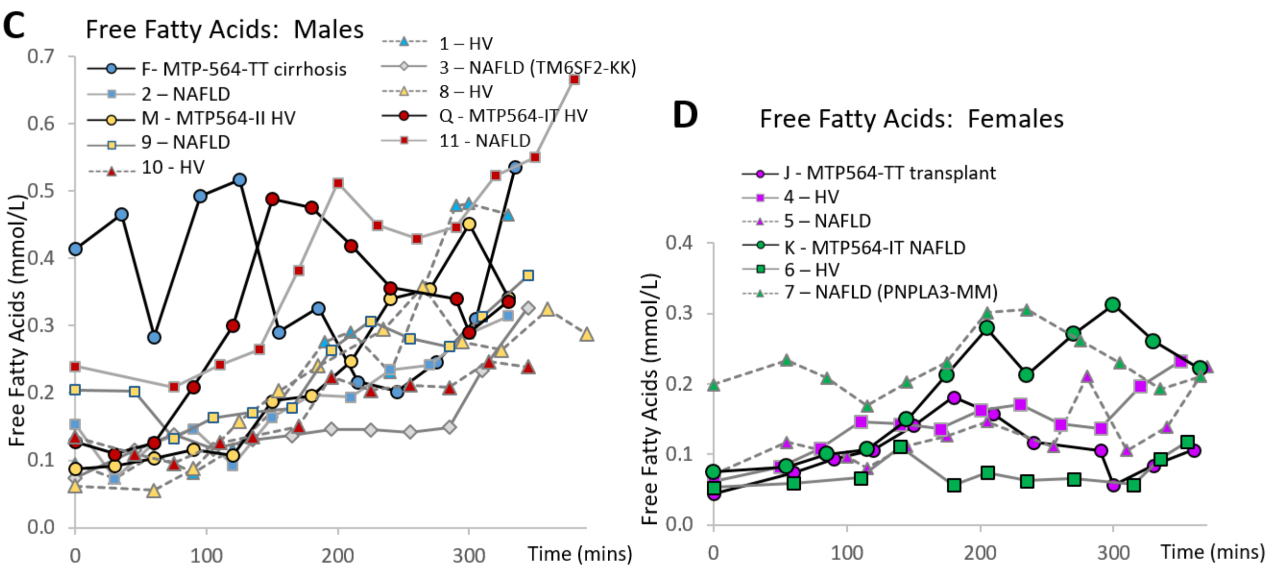


**Supplementary Figure 1. Serum cholesterol levels and free fatty acid levels in study participants.** Participants grouped according to age and gender matching with family member (Fig. 1a and Supplemental Table 3). Genes are shown in parentheses where participant is homozygous for other effect alleles: *PNPLA3* rs738409; *TM6SF2* rs58542926). **A** Fasting cholesterol. **B** Cholesterol level approx. 2h after eating standard study meal. **C** Free fatty acid levels in males. **D** Free fatty acid levels in females. NAFLD= non-alcoholic fatty liver disease; HV=healthy volunteer; FL=fatty liver. *PNPLA3* (rs738409) variant homozygotes (148-MM) and *TM6SF2* (rs58542926) variant homozygotes (128-KK) are indicated.

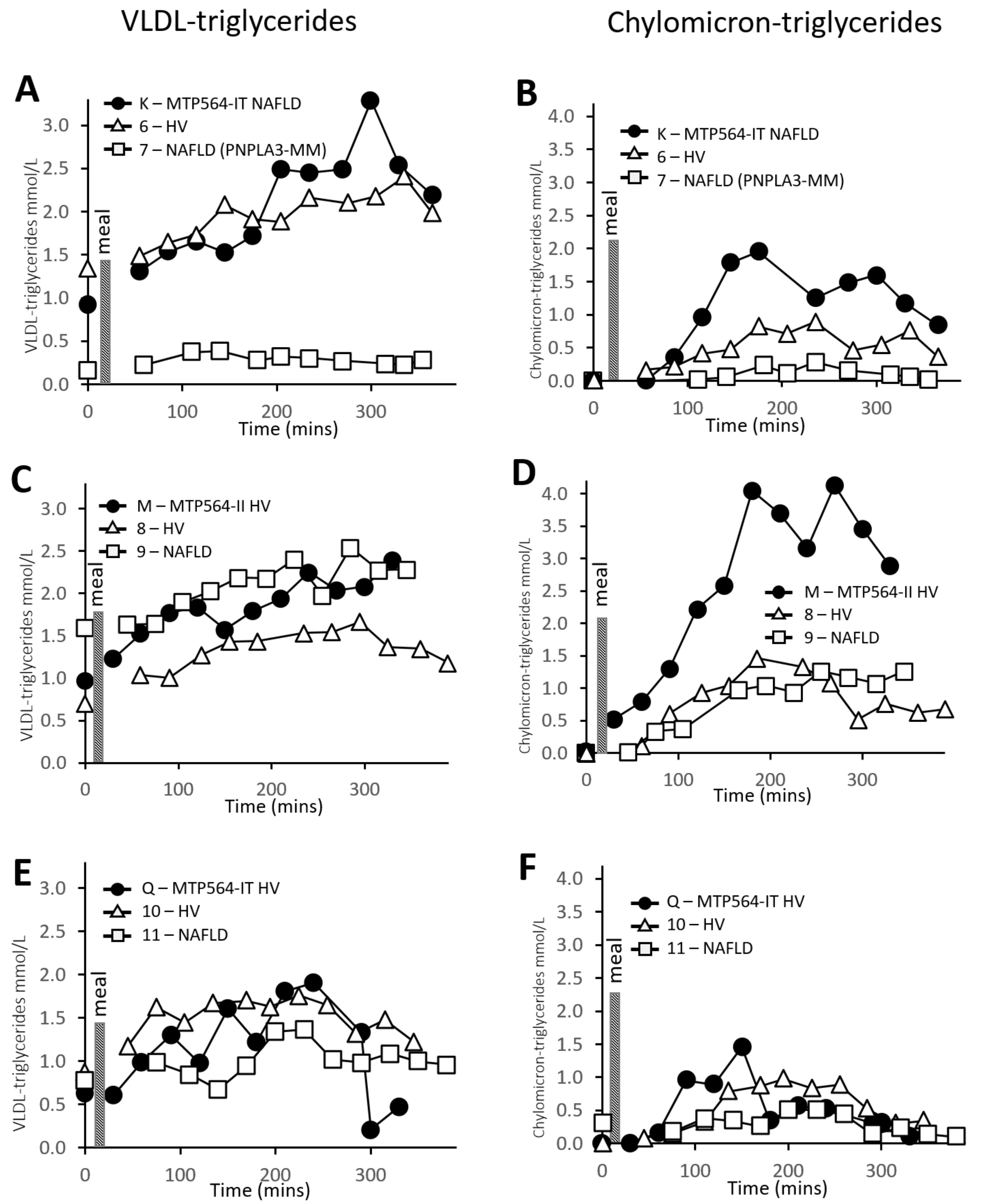


**Supplementary Figure 2. Lipoprotein-associated triglyceride levels in study participants before and after a fatty meal.** Participants are described in Supplemental Table 1**. A** Serum VLDL-triglycerides in MTP564-IT family member K and matched control participants: 6 (HV) and 7 (NAFLD patient). **B** Chylomicron-triglycerides in participants K, 6 and 7. **C** VLDL-triglyceride in MTP564-II (wild type) family member M and matched control participants: 8 (HV) and 9 (NAFLD patient). **D** Chylomicron-triglycerides in participants M, 8 and 9. **E** VLDL-triglycerides in MTP564-IT family member Q and matched control participants: 10 (HV) and 11 (NAFLD patient). **F** Chylomicron-triglycerides in participants Q, 10 and 11. *PNPLA3* (rs738409) variant homozygote (148-MM) is indicated. HV=healthy volunteer; NAFLD=non-alcoholic fatty liver disease.

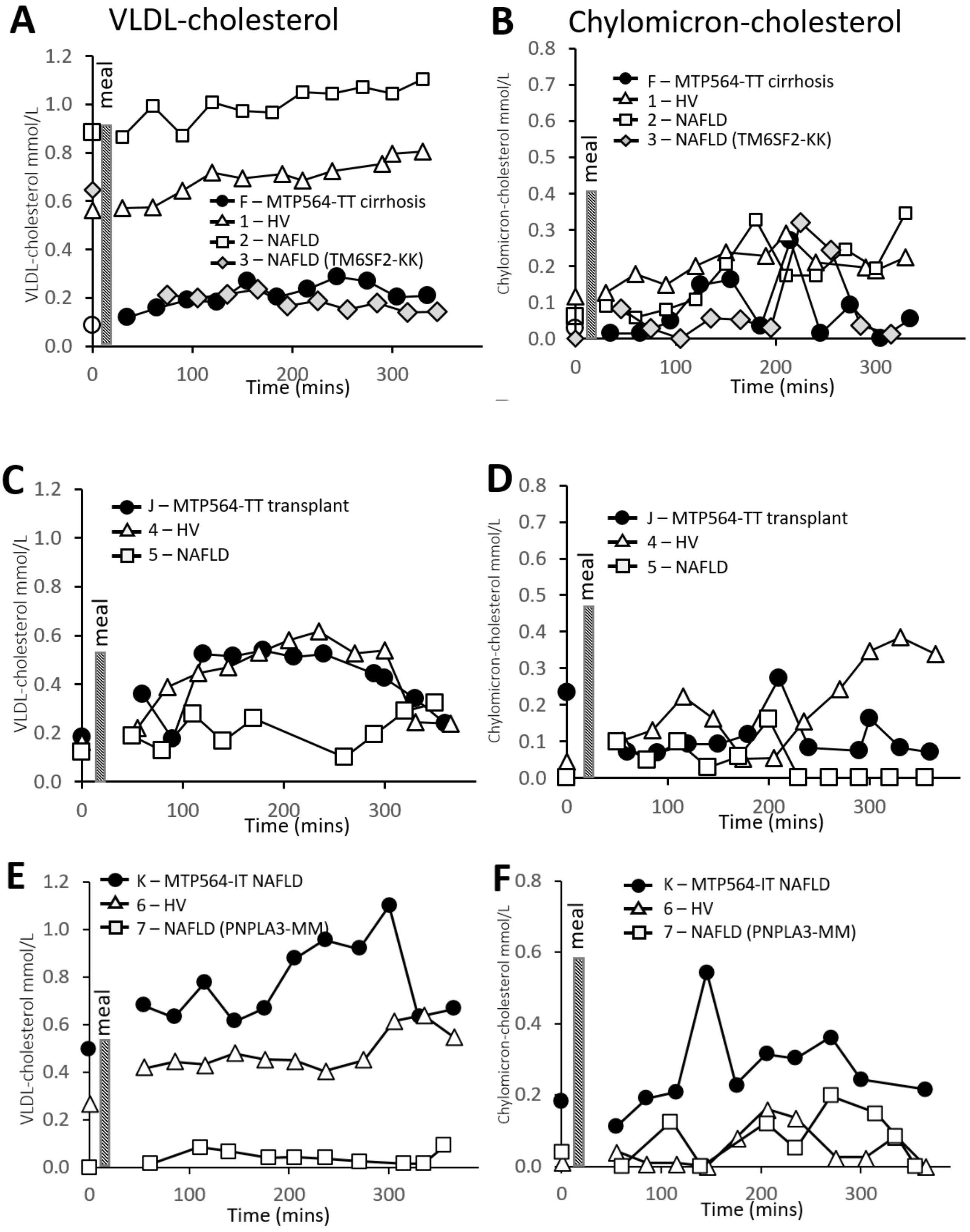
A
VLDL-cholesterol
VLDL-cholesterol mmol/L
meal
F – MTP564-TT cirrhosis
1 – HV
2 – NAFLD
3 – NAFLD (TM6SF2-KK)
Time (mins)
B
Chylomicron-cholesterol
Chylomicron-cholesterol mmol/L
F – MTP564-TT cirrhosis
1 – HV
2 – NAFLD
3 – NAFLD (TM6SF2-KK)
C
J – MTP564-TT transplant
4 – HV
5 – NAFLD
D
J – MTP564-TT transplant
4 – HV
5 – NAFLD
E
K – MTP564-IT NAFLD
6 – HV
7 – NAFLD (PNPLA3-MM)
F
K – MTP564-IT NAFLD
6 – HV
7 – NAFLD (PNPLA3-MM)

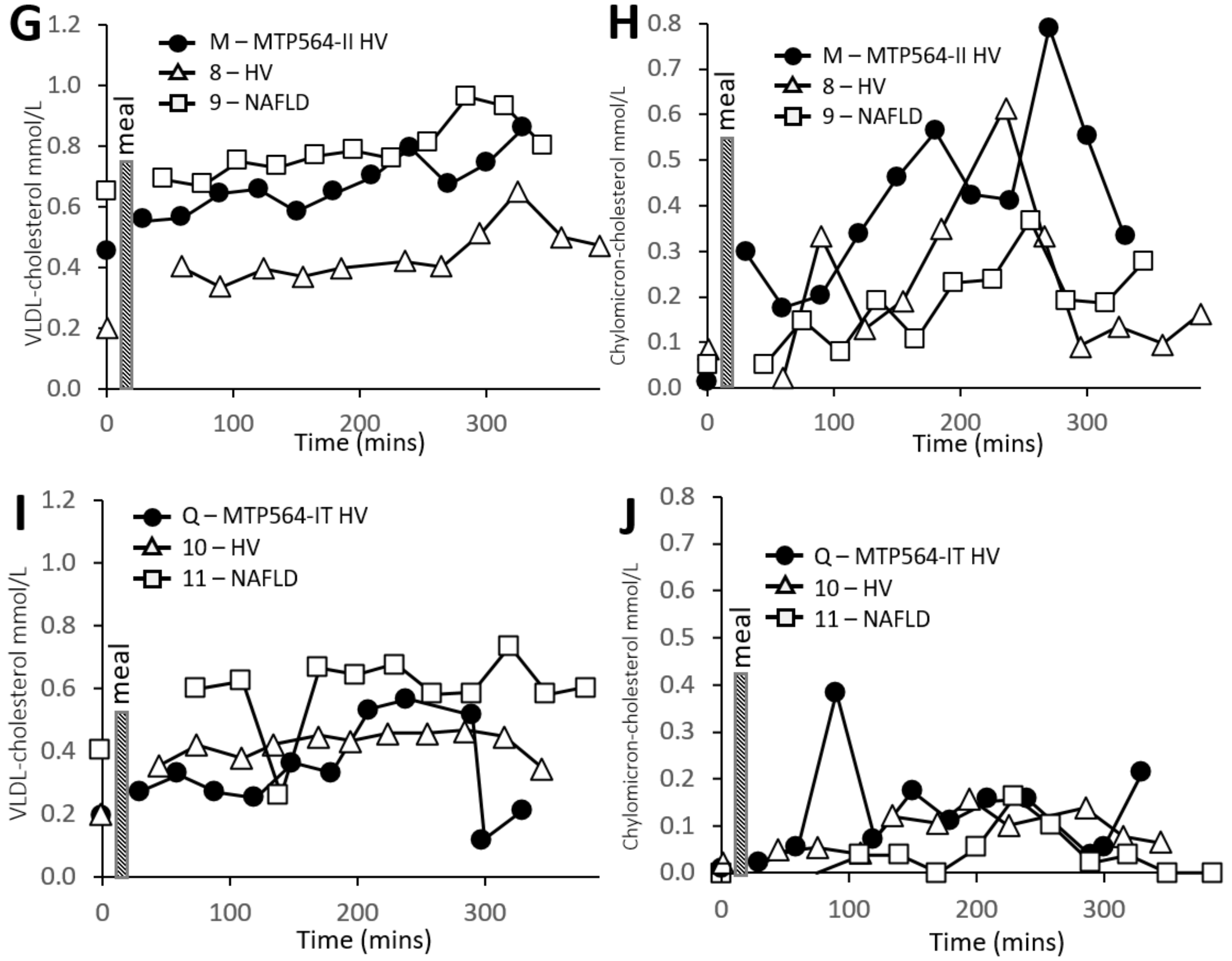


**Supplementary Figure 3. Lipoprotein-associated cholesterol levels in study participants before and after a high fat meal.** Participants are described in Supplemental Table 1. **A** VLDL-associated cholesterol levels in MTP564-TT family member F with cirrhosis and matched control participants: 1 (HV), 2 and 3 (NAFLD patients). **B** Chylomicron**-**associated cholesterol in participants F, 1, 2 and 3. **C** VLDL-cholesterol in MTP564-TT family member J with liver transplantand matched control participants: 4 (HV) and 5 (NAFLD patient). **D** Chylomicron**-**associated cholesterol in participants J, 4 and 5. **E** VLDL-associated cholesterol levels in MTP564-IT family member K with NAFLD and matched control participants: 6 (HV) and 7 (NAFLD patient). **F** Chylomicron**-**associated cholesterol in participants K, 6 and 7. **G** VLDL-associated cholesterol levels in MTP564-II (wild type) family member M and matched control participants 8 (HV) and 9 (NAFLD patient). **H** Chylomicron**-**associated cholesterol in participants M, 8 and 9. **I** VLDL-associated cholesterol levels in MTP564-IT family member Q and matched control participants 10 (HV) and 11 (NAFLD patient). J Chylomicron**-**associated cholesterol in participants Q, 10 and 11.

*TM6SF2* (rs58542926) variant homozygote (128-KK) and *PNPLA3* (rs738409) variant homozygotes (148-MM) are indicated. HV=healthy volunteer; NAFLD=non-alcoholic fatty liver disease.

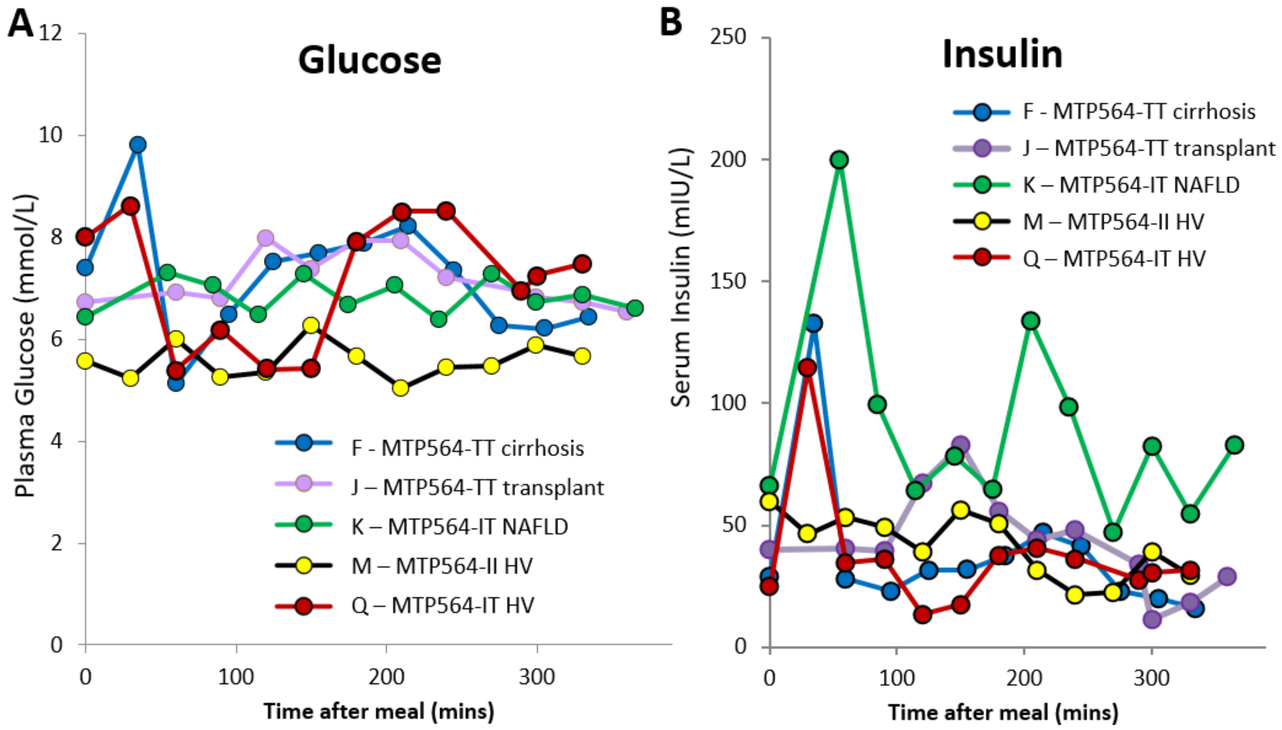


**Supplementary Figure 4 Blood biomarker levels in family members after consuming a fatty meal**. **A Fasting blood glucose. B Insulin levels.**

*PNPLA3* (rs738409) variant homozygotes (148-MM) and *TM6SF2* (rs58542926) variant homozygotes (128-KK) are indicated. HV=healthy volunteer; NAFLD=non-alcoholic fatty liver disease.

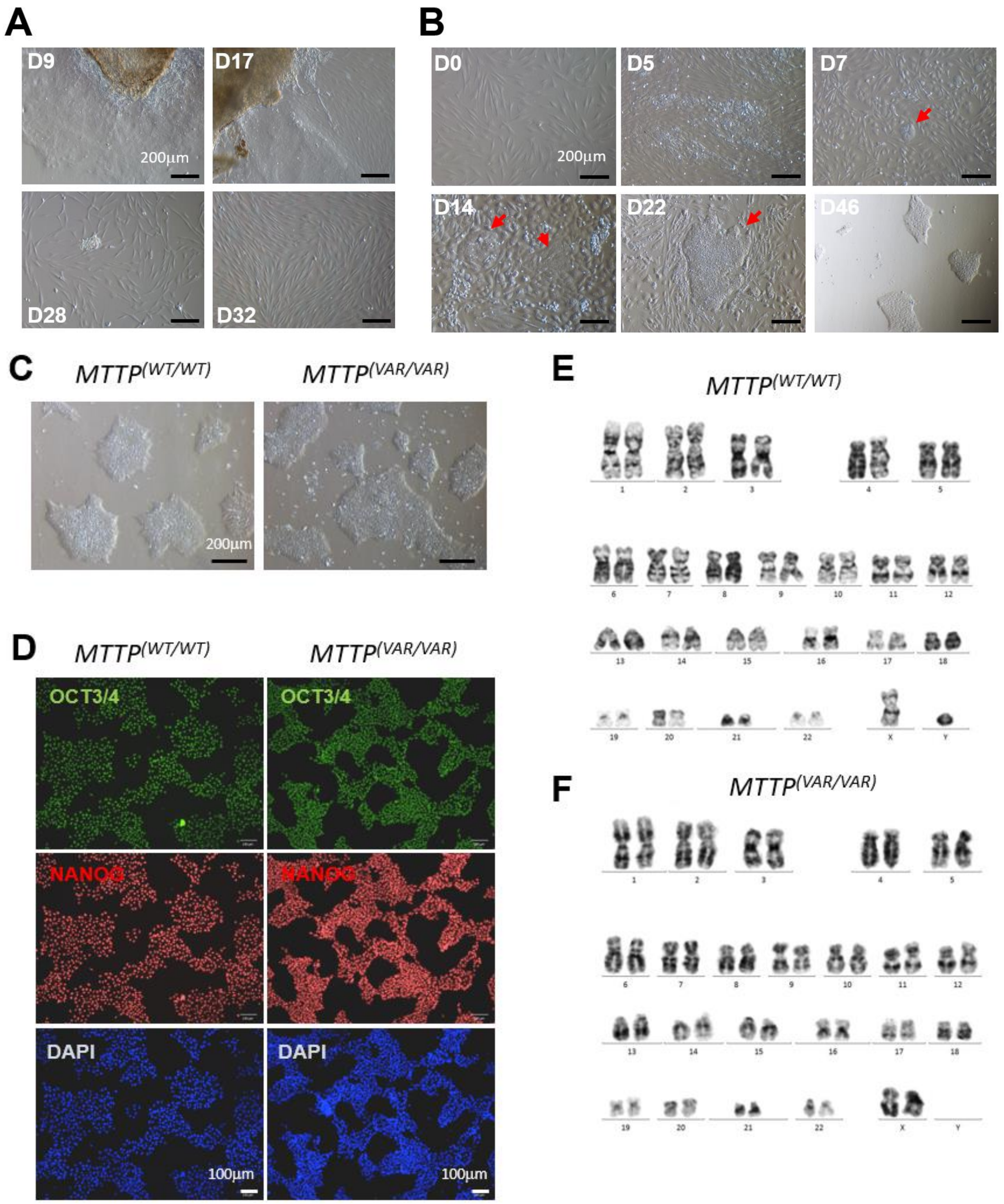


**Supplementary Figure 5. Characteristics of hiPSCs derived from donor skin biopsy**. **a**, Light microscopy showing representative skin biopsy fibroblast outgrowth at day 9, 17, 28 and 32. **b**, Light microscopy showing representative images of fibroblast reprogramming at day 0, 5, 7, 14, 22 and 46 post viral transduction. Red arrows indicate emerging hIPSC colonies. **c**, Light microscopy showing representative pictures of reprogrammed *MTTP*$^{(WT/WT)}$ and *MTTP*$^{(VAR/VAR)}$ hIPSC cultures. **d**, Immunocytochemistry showing expression of pluripotency markers in *MTTP*$^{(WT/WT)}$ and *MTTP*$^{(VAR/VAR)}$ hiPSCs. **e**, Karyotype of *MTTP*$^{(WT/WT)}$. **f,** Karyotype of *MTTP*$^{(VAR/VAR)}$ hiPSCs.

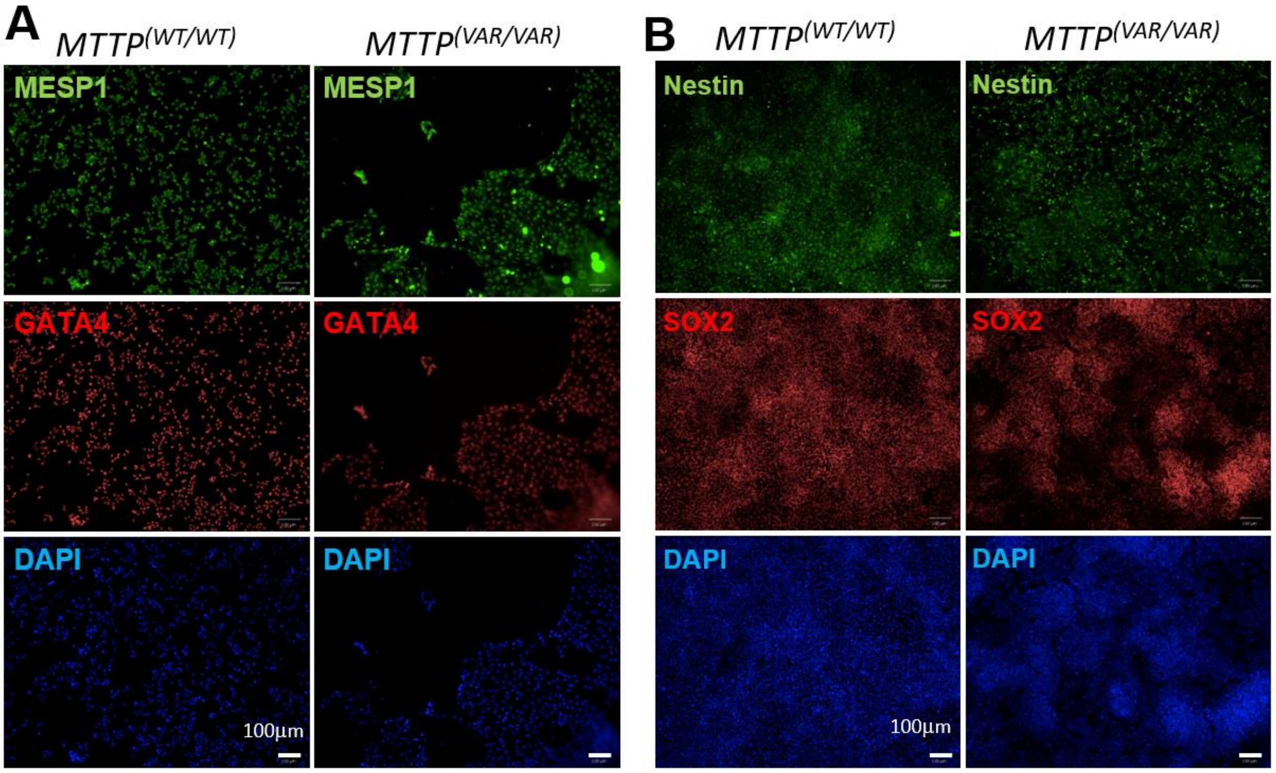


**Supplementary Figure 6. Differentiation of *MTTP*$^{(WT/WT)}$ and *MTTP*$^{(VAR/VAR)}$ hiPSCs into multiple germ layers. A** Expression of mesodermal genes following mesoderm differentiation of *MTTP*$^{(WT/WT)}$ and *MTTP*$^{(VAR/VAR)}$ hiPSCs by immunocytochemistry with MESP1, GATA4 and DAPI staining. **B** Expression of ectoderm genes following ectoderm differentiation of *MTTP*$^{(WT/WT)}$ and *MTTP*$^{(VAR/VAR)}$ hiPSCs by immunocytochemistry with Nestin, SOX2 and DAPI staining.

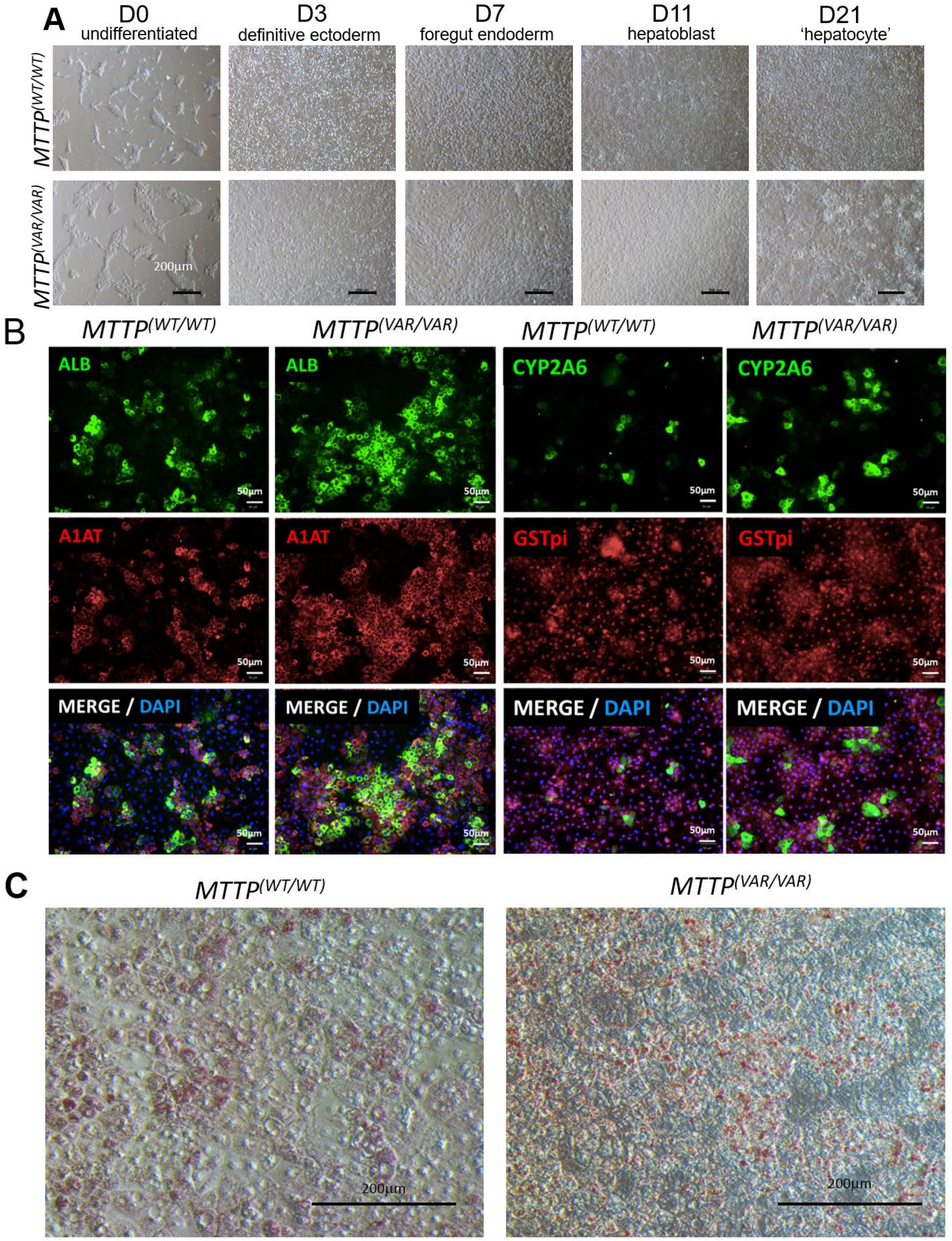


**Supplementary Figure 7. Differentiation of *MTTP*$^{(WT/WT)}$ and *MTTP*$^{(VAR/VAR)}$ hiPSCs into mature HLCs. A** Representative light microscopy images of *MTTP*$^{(WT/WT)}$ and *MTTP*$^{(VAR/VAR)}$ hiPSCs as they differentiate into HLCs including undifferentiated cells (Day 0), definitive endoderm (Day 3), foregut endoderm (Day 7), hepatoblast (Day 11) and 'hepatocytes' (Day 21). **B** Immunocytochemistry showing expression of hepatocyte markers in *MTTP*$^{(WT/WT)}$ and *MTTP*$^{(VAR/VAR)}$ hiPSC derived HLCs. **C** Light microscopy of Oil-Red-O stained *MTTP*$^{(WT/WT)}$ and *MTTP*$^{(VAR/VAR)}$ HLCs.
